# Genetic Heterogeneity and Homogeneity Among Orofacial Cleft Subtypes: Genome-Wide Association Studies in the Cleft Collective

**DOI:** 10.1101/2025.05.02.25326852

**Authors:** Kyle Dack, Kerstin U. Ludwig, Evie Stergiakouli, Jonathan Sandy, Sethlina Aryee, George Davey Smith, Amy Davies, Yvonne Wren, Gemma C Sharp, Kerry Humphries, Elisabeth Mangold, Lucy Goudswaard, Karen Ho, Tom Dudding, Sarah J. Lewis

## Abstract

Several genome wide association studies (GWASs) of orofacial cleft have been conducted, however the majority of studies to date have restricted their analysis to non-syndromic cleft lip with/without cleft palate and have not analysed all orofacial clefts combined, or conversely, investigated heterogeneity between subtypes. We conducted a GWAS of orofacial clefts within 2,268 cases from the Cleft Collective and 7,913 population-based controls; we performed analyses of all orofacial clefts, plus 7 different subgroups. We replicated our findings in a meta-analysis of independent samples and investigated patterns of correlation across subgroups. We identified 27 regions at genome-wide significance, 8 of which were novel. Although our sample size was small (n cases=237) we included the first GWAS of Pierre Robin Sequence, we found one genome wide significant SNP (P<5x10^-8^), and another 23 suggestive associations (P<10^-5^). Novel loci include those mapping to *LHX8* (combined clefts), *ARHGEF18* and *ARHGEF19* (cleft lip with/without palate), *FBN2* (cleft lip only), *SLC35B3* (cleft palate only), *CASC20* (Pierre Robin Sequence) and *CHRM2* (non-syndromic cleft palate only). Several novel hits were in regions previously associated with facial morphology in GWAS or were in regions involved in key developmental processes, including neural crest cell migration and craniofacial development. We identified genetic loci with similar effects across all subgroups and some loci which were subtype specific, we also identified 3 loci with opposing effects on cleft lip and Pierre Robin sequence. Our findings highlight the merit of including all orofacial cleft subtypes in GWAS studies, and investigating heterogeneity of effects across subtypes.

## Introduction

Orofacial clefts are among the most common congenital birth anomalies, occurring in approximately 1 in 700 births globally (Panamonta et al., 2015). These anomalies can manifest as cleft lip only, cleft palate only, or a combination of both cleft lip and palate (Mossey et al., 2009). Orofacial clefts occur in syndromic or non-syndromic forms, with non-syndromic clefts being more prevalent than syndromic (Mossey et al., 2009 and Davies et al., 2024). Syndromic clefts typically occur alongside other anomalies or health issues and are usually caused by a rare single gene or chromosomal mutation, although the penetrance of these mutations can differ between individuals resulting in heterogeneity of the phenotype (Leslie and Marazita, 2013). In contrast, non-syndromic clefts follow a multifactorial aetiology and, accordingly, are influenced by both genetic and environmental factors (Dixon et al., 2011).

Pierre Robin sequence (PRS) presents as a triad of anomalies arising from a single underlying cause. The three primary characteristics include micrognathia (a small and retrusive mandible), leading to glossoptosis (a downwardly displaced or retracted tongue), which subsequently obstructs the upper airway and causes breathing difficulties. Additionally, a wide, U-shaped cleft palate is frequently present (Gangopadhyay et al., 2012). Rare functional mutations in the *SOX9* gene on chromosome 17 are established risk factors for PRS, though environmental factors and additional genes also play a role (Selvi et al., 2013 and Varadarajan et al., 2021).

Genome wide association studies (GWAS) of orofacial clefts have identified over 50 genetic risk loci associated with non-syndromic cleft lip with or without cleft palate (nsCL/P) but relatively few loci have been identified for cleft palate only (CPO) (Mangold et al, 2010; Beaty et al, 2010; Ludwig et al, 2012; Yu et al, 2017; Leslie et al, 2017; Butali et al, 2019; Birnbaum et al. 2009; Welzenbach et al. 2021 and Mukhopadhyay et al, 2021). To our knowledge there haven’t been any GWAS of Pierre Robin Sequence to date.

Syndromic forms of cleft are typically excluded from GWAS as these are thought to add noise to the analysis due to having a monogenic cause. However, recent evidence suggests that the boundaries between syndromic and non-syndromic clefts, as well as between different cleft subtypes, are less distinct than previously assumed (Wilson et al., 2023). Both rare and common genetic variants appear to contribute to all cleft subtypes (Wilson et al., 2023). Exome sequencing studies have identified single pathogenic mutations in non-syndromic orofacial cleft families (Demeer et al., 2019). In addition, common genetic variants have been shown to contribute to the variation of neurodevelopmental conditions which were thought to have a monogenic cause (Huang et al., 2024), such variants may well contribute to syndromic orofacial clefts.

Furthermore, genetic variants identified at genome-wide significance in one cleft subtype might also influence other subtypes, albeit with smaller effect sizes; if this is the case combining genetic data across subtypes may increase the power to detect new loci. Although, a previous study did combine cleft lip and cleft palate only subtypes (Leslie et al., 2016), the study did not include syndromic cases and studies have yet to systematically evaluate the magnitude of effects of SNPs across multiple cleft subtypes.

The United Kingdom (UK) based Cleft Collective (Davies et al, 2024), at the University of Bristol, is one of the largest cohort studies of children born with cleft and their families. The aim of this study was to conduct GWASs to identify new genetic variants for all orofacial clefts and its subtypes and to investigate the heterogeneity between subtypes using a case-control design.

## Results

After applying quality control procedures, we were left with 2,268 cases and 7,913 controls of European ethnicity for our GWAS analyses. Figure 1 shows the number individuals excluded at each stage of the QC process and the final numbers of each subtype included in our analyses.

**Figure 1.**
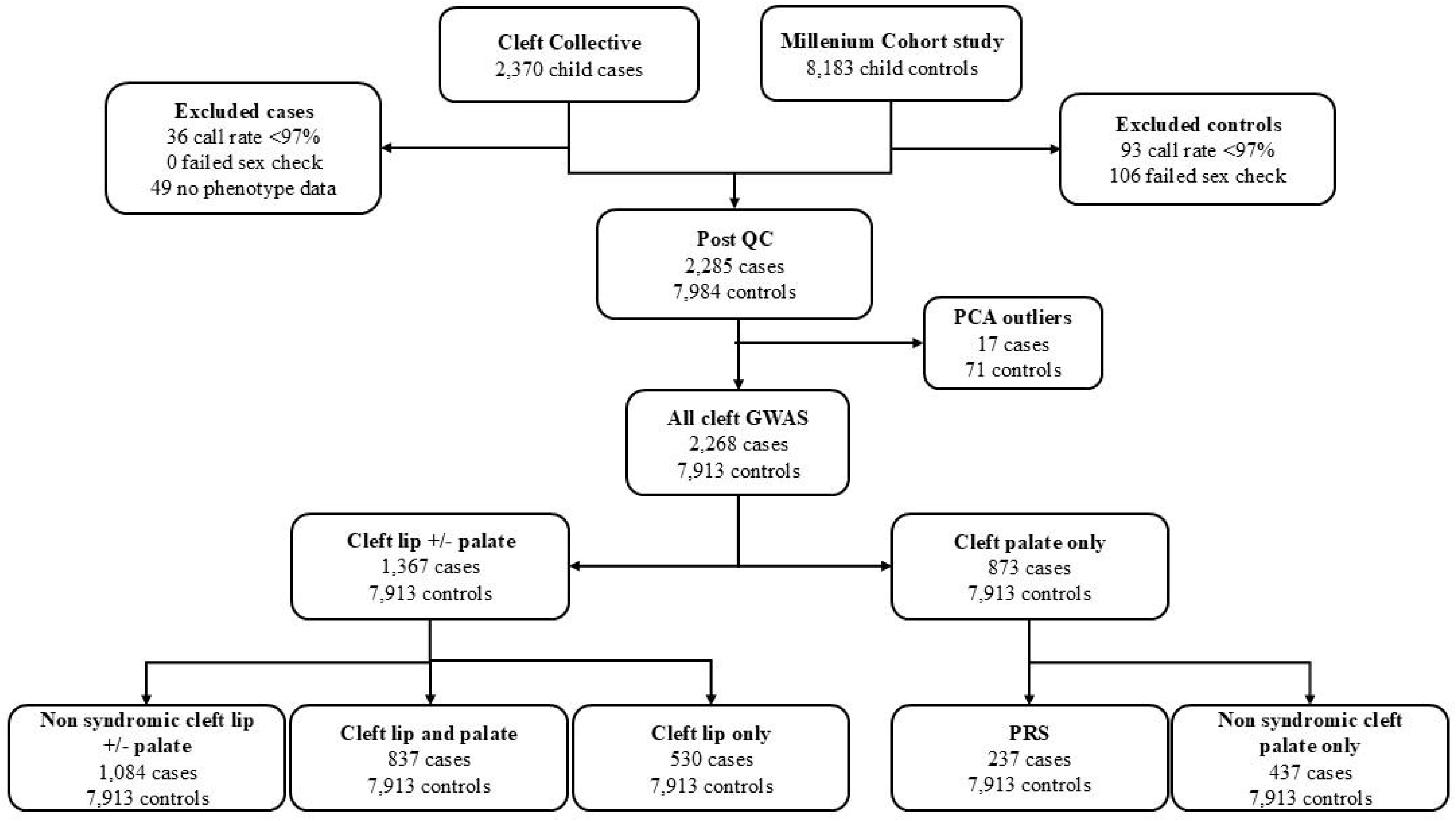
Flow chart showing number of individuals included in our combined cases and subgroup analyses following quality control exclusions

Lambda values for GWAS inflation ranged from 1.02 to 1.13 in our GWAS analyses (Supplementary Table S1). We identified 1,077 genome-wide-significant SNPs (p<5x10^-8^), which reduced to 27 lead SNPs after clumping (Table 2). Of the regions identified, 19 had been reported in previous orofacial cleft studies (indicated by the GWAS catalogue), while 8 were novel (Table 3). Our strongest association was between a previously identified SNP (rs17242358) on chromosome 8q24 and nsCL/P (p=3.36x10^-42^). Supplementary Table S2 shows a summary of the number of genome wide significant associations by cleft subtype and Supplementary Table S3 shows odds ratios and p-values for our lead SNPs across all cleft subtypes.

**Table 1.** Description and acronyms of orofacial cleft subtypes.

|  |  |  |
| --- | --- | --- |
| **Cleft lip with or without cleft palate** | CL/P | Children born with a cleft lip regardless of whether they also had a cleft palate or not and regardless of whether they had a syndrome or not |
| **non-syndromic cleft lip with or without cleft palate** | nsCL/P | As for CL/P above but excluding any children known to have a syndrome |
| **Cleft lip with cleft palate** | CLP | Children born with a cleft lip plus a cleft palate whether or not they are known to have a syndrome |
| **Cleft lip only** | CLO | Children born with a cleft lip but not a cleft palate, includes children with and without a syndrome or sequence. |
| **Cleft palate only** | CPO | Children born with a cleft palate but not a cleft lip, includes those with and without a syndrome or sequence (e.g. Pierre Robin sequence (PRS)) |
| **non-syndromic cleft palate only** | nsCPO | Children with a cleft palate but not a cleft lip, excluding children with a known syndrome or sequence |
| **Pierre-Robin Sequence** | PRS | Children who have been diagnosed as have PRS regardless of whether they also have a syndrome or not |

**Table 2.** Lead genome-wide significant SNPs.

| Lead SNP | Location (GRCh37) | Chromosome band | Mapped gene/s | chromosomal range of GWAS sign SNPs | A 1 | A2 | MAF controls | 95% credible interval | Lead phenotype | Phenotypes with $p < 10^{-4}$ | Lead phenotype OR | P-value | Replicates ? |
| --- | --- | --- | --- | --- | --- | --- | --- | --- | --- | --- | --- | --- | --- |
| rs61769781 | 1:16544626 | 1p36.13 | ARHGEF19 | 16537366-1655264 | A | G | 0.11 | 16514898-16633123 | nsCL/P | All lip phenotypes | 1.44 | 1.81E-8 | Yes, P=0.0014 |
| rs9439714 | 1:18976489 | 1p36.13 | PAX7* (intron) | 18920364-19027239 | C | T | 0.35 | 18965061-18978372 | CL/P | All lip phenotypes | 1.45 | 4.06E-18 | Yes, P=1.1E-16 |
| rs4112328 | 1:75500597 | 1p31.1 | LHX8 | 75498339-75589337 | A | G | 0.43 | 75498339-75577086 | All cleft | CL/P nsCL/P CLP<sup>1</sup> | 1.22 | 3.73E-09 | No, p=0.94 |
| rs4147827 | 1:94548080 | 1p22.1 | ABCA4* (intron) | 94540675-94558110 | G | C | 0.18 | 94548080-94558110 | CL/P | All lip phenotypes | 1.36 | 7.49E-10 | Yes, p=1.43E-14 |
| rs126280 | 1:210019824 | 1q32.2 | UTP25* (intron) | 210019824-210020013 | A | G | 0.23 | 209950760-210022893 | CL/P | nsCL/P CLO<sup>2</sup> | 1.49 | 1.86E-18 | Yes, p=8.52E-14 |
| rs10172734 | 2:16733054 | 2p24.2 | CYRIA* (3' UTR) | 16634551-16774094 | A | G | 0.3 | 16706891-16733054 | CL/P | All lip phenotypes | 1.40 | 1.58E-14 | Yes, p=8.89E-08 |
| rs4952552 | 2:42198217 | 2q14.2 | PKDCC / LINCO2898 | 42181679-42213484 | A | G | 0.47 | 42179157-42215172 | CL/P | All Cleft nsCL/P CLP<sup>2</sup> | 0.77 | 2.66E-09 | Yes, p=0.0074 |
| rs6741434 | 2:43695148 | 2p21 | THADA* (downstream transcript) | 43540125-43741440 | T | C | 0.24 | 43540125-43741440 | NSCL/P | CL/P CLO | 1.36 | 2.45E-09 | Yes, p=0.0015 |
| rs79482068 | 3:189541869 | 3q28 | TP63* (intron) | 189537013-189553372 | T | C | 0.06 | 189541869-189547735 | CL/P | All lip phenotypes | 1.69 | 2.97E-13 | Yes, p=5.58E-05 |
| rs1347188 | 4:124743259 | 4q28.1 | LINC01091* (intron) | 124743259-12474571 | G | A | 0.24 | 124678521-124766956 | CLP | CL/P nsCL/P | 1.37 | 2.58E-08 | Yes, p=0.0171 |
| rs62390705 | 5:127913096 | 5q23.3 | FBN2 | 127911108-127914018 | G | A | 0.13 |  | CLO | CL/P nsCL/P<sup>2</sup> | 1.58 | 4.00E-08 | Yes, p=0.0088 |
| rs72823645 | 6:8359186 | 6p24.3 | SLC35B3 | 8359186-8359186 | T | C | 0.05 | 8359186-8359186 | CPO | All cleft nsCPO | 1.69 | 3.67E-08 | No CPO |
| rs28361060 | 6:32303848 | 6p21.32 | TSBP1* (intron) | 32273060-32303848 | A | G | 0.22 | 32212655-32338386 | All cleft | All lip phenotypes | 0.75 | 7.97E-09 | No, p=0.1524 |
| rs17168736 | 7:13630495 | 7q33 | CHRM2 | 136304959- | A | C | 0.18 | 136296025- | nsCPO | All clefts | 1.56 | 3.35E-08 | No CPO |
|  | **9** |  |  | **136304959** |  |  |  | **136321655** |  | **CPO** |  |  |  |
| rs12543318 | 8:88868340 | 8q21.3 | DCAF4L2 | 88868340-89017568 | C | A | 0.35 | 88868340-89017568 | CL/P | All lip phenotypes | 1.30 | 5.67E-10 | Yes, p=3.09E-11 |
| rs17242358 | 8:129964873 | 8q24.21 | CCDC26/LINC00976 | 128194706-130023578 | A | G | 0.18 | 129929311-129990382 | CL/P | All lip phenotypes | 1.92 | 3.36E-42 | Yes, p=1.2E-62 |
| rs7870795 | 9:100574120 | 9q22.33 | FOXE1/PTCSC2* | 100573164-100658123 | T | C | 0.29 | 1005753164-100620730 | All cleft | All lip phenotypes<sup>1</sup> | 0.77 | 3.78E-11 | Yes, p=9.28E-06 |
| rs1898349 | 10:118817067 | 10q25.3 | SHTN1* (upstream transcript) | 118786985-118869886 | T | C | 0.17 | 118786985-118869886 | CL/P | All cleft nsCL/P CL/P<sup>3</sup> | 1.37 | 1.40E-09 | Yes, p=8.31E-10 |
| rs9601323 | 13:80700397 | 13q31.1 | SPRY2 | 80679302-80702065 | A | G | 0.46 | 80689877-80701485 | CLP | All cleft nsCL/P CL/P<sup>3</sup> | 1.46 | 1.01E-12 | Yes, p=3.32E-11 |
| rs10483604 | 14:51485195 | 14q22.1 | TRIM9* (intron) | 51427267-51490767 | T | C | 0.23 | 51427267-51490767 | CL/P | All cleft nsCL/P CL/P<sup>3</sup> | 1.33 | 1.37E-09 | Yes, p=0.0143 |
| rs4901118 | 14:51856109 | 14q22.1 | FRMD6 | 51856109-51856566 | G | A | 0.39 | 51856109-51856566 | CL/P | nsCL/P CLP CLO | 0.77 | 1.51E-09 | Yes, p=1.58E-08 |
| rs2600519 | 15:33063809 | 15q13.3 | FMN1* | 33058523-33064636 | G | A | 0.34 | 33058304-33064636 | CL/P | All cleft nsCL/P CLP<sup>3</sup> | 0.75 | 7.40E-10 | Yes, p=3.42E-06 |
| rs8044196 | 16:3984034 | 16p13.3 | CREBBP_AD CY9* | 3918004-3997030 | A | G | 0.33 | 3918004-3985081 | CL/P | All cleft lip phenotypes | 1.33 | 4.58E-11 | Yes, p=0.0007 |
| **rs12971753** | **19:7499238** | **19p13.2** | **ARHGEF18\* (intron)** | **7447672-7529415** | **C** | **T** | **0.23** | **7447672-7529415** | **CL/P** | **All cleft nsCL/P CLP<sup>3</sup>** | **0.73** | **3.42E-09** | **No, p=0.082 (same direction)** |
| **rs2225351** | **20:6484640** | **20p12.3** | **CASC20\* (intron)** | **6428764-6591010** | **T** | **C** | **0.42** | **6428764-6591010** | **CPO** | **PRS<sup>4</sup>** | **1.33** | **1.16E-08** | **No, CPO** |
| rs34753522 | 20:39278391 | 20q12 | MAFB | 39268516-39278391 | C | T | 0.36 | 39268516-39278391 | CL/P | All cleft nsCL/P CLP<sup>3</sup> | 0.76 | 6.84E-10 | Yes, p=0.0056 |
| rs6018099 | 20:45473014 | 20q13.12 | RN7SKP33* | 45440006-45486817 | G | A | 0.48 | 45440006-45486817 | CL/P | All cleft nsCL/P | 1.30 | 6.32E-10 | Yes, p=0.0018 |
|  |  |  |  |  |  |  |  |  |  | CLP<sup>3</sup> |  |  |  |

A1= minor and effect allele, A2= reference allele,
\*= falls within the gene region
lead phenotype=phenotype which has the smallest p-value for association with the lead phenotype
Mapped gene is either the one which the SNP falls within or the nearest gene to the variant according to dbSNP (https://www.ncbi.nlm.nih.gov/SNP)
<sup>1</sup>Nominally associated with all phenotypes p<0.05,
<sup>2</sup>Effect in opposite direction for PRS, p<0.05
<sup>3</sup>Also nominally associated with CLO p<0.05
<sup>4</sup>Effect not present in nsCPO, greatest effect size in PRS only cases OR=1.7

**Table 3.** Evidence linking GWAS significant regions from case-control analyses to orofacial cleft.

| rsID | CHR | Chromosome location (GRCh37) | Nearest gene | Prior evidence from GWAS |  |  | Gene function | Other evidence linking gene to cleft | References |
| --- | --- | --- | --- | --- | --- | --- | --- | --- | --- |
|  |  |  |  | SNP | Gene | Region |  |  |  |
| rs61769781 | 1 | 16544626 | ARHGEF19 | N | N | N | *Guanine nucleotide exchange factor (GEF) for RhoA GTPase. Regulation of cell shape, small GTPase mediated signal transduction* | Gene previously associated with facial morphology (right endocanthion-left cheilion distance). | Xiong et al, 2019 |
| rs9439714 | 1 | 18976489 | PAX7 | Y | Y | Y | *Paired box (PAX) family of transcription factor critical role during fetal development and cancer growth.* |  | Leslie et al, 2017<br>Ray D et al 2021 |
| rs4112328 | 1 | 75500597 | LHX8 | N | N | N | *Transcription factor that plays a role in patterning and differentiation of tissues including tooth morphogenesis.* | Disruption of *LHX8* gene function caused impairment in palatal shelf contact and fusion, leading to formation of a cleft secondary palate.<br>Vieira sequenced DNA from CLP patients and found a mutation for *LHX8* | Zhao et al, 1999<br>Vieira et al, 2005 |
| rs4147827 | 1 | 94548080 | ABCA4 | N | Y | Y | *Transport of molecules across extra- and intracellular membranes – retina specific* | Nearest GWAS SNP rs3789432 at 1:94575308 | Beaty et al, 2010<br>Leslie et al, 2017<br>Yu et al 2017 |
| rs126280 | 1 | 210019824 | UTP25 (IRF6) | N | Y | Y | *Component of ribosome, enables RNA binding activity.* | *Just upstream of IRF6*<br>Nearest GWAS SNP rs2064163, 1:210048819. | Yu et al 2017<br>Beaty et al, 2010<br>Leslie et al, 2017<br>Huang et al, 2019 |
| rs10172734 | 2 | 16733054 | CYRIA | Y | Y | Y | *Predicted to enable small GTPase binding activity and involved in regulation of actin filament polymerization.* |  | Yu et al, 2017<br>Leslie et al, 2019<br>Mukhapadhyay et al, 2021<br>Yang et al, 2020 |
| rs4952552 | 2 | 42198217 | PKDCC<br>LINC02898 | N | Y | Y | *Secreted tyrosine-protein kinase that mediates phosphorylation of extracellular proteins and endogenous proteins in the secretory pathway.* | Previously associated with cleft parent endophenotype (Indencleef et al, 2021). Imuta et al. (2009) generated Pkdcc -/- mice, resulted in shortening of the long bones of the limbs and cleft palate. | Ludwig et al, 2017<br>Leslie et al, 2017<br>Indencleef et al, 2021<br>Imuta et al, 2019 |
| rs6741434 | 2 | 43695148 | THADA | N | Y | Y | *Methylates the 2'-O-ribose of nucleotides at position 32 of the anticodon loop of substrate tRNAs.* |  | Mangold et al, 2010<br>Ludwig et al, 2012 |
| rs79482068 | 3 | 189541869 | TP63 | N | Y | Y | *Transcription factor, Involved in NOTCH signalling by probably inducing JAG1 and JAG2. Plays a role in the regulation of epithelial morphogenesis.* | Nearest GWAS SNP rs76479869, 3:189553372 (also sign in our analysis). | Leslie et al 2017 |
| rs1347188 | 4 | 124743259 | LINC01091 | N | Y | Y | *Long Intergenic Non-Protein Coding RNA 1091.* |  | Yu et al, 2017<br>Butali et al, 2019 |
| rs62390705 | 5 | 127913096 | FBN2 | N | N | N | *Component of connective tissue microfibrils and may be involved in elastic fibre assembly.* | SNPs at position 128107907 and 128190363 have been found to be associated with lip morphology. | White et al, 2020<br>Hoskins et al, 2021 |
| rs72823645 | 6 | 8359186 | SLC35B3 | N | N | N | *Solute Carrier Family 35 (Adenosine 3'-Phospho 5'-Phosphosulfate Transporter) gene.* | Related pathway involved in chondrodysplasia with Joint Dislocations, Gpapp Type (CDP-GPAPP), disorder characterised by short stature, chondrodysplasia with brachydactyly, congenital joint dislocations, cleft palate, and facial dysmorphism. | Paganini et al, 2020 |
| rs28361060 | 6 | 32303848 | TSBP1 | N | N | N | *Testis expressed basic protein. Associated with connective tissue diseases.* | Just upstream of *NOTCH4* involved in craniofacial development. |  |
| rs17168736 | 7 | 136304959 | CHRM2 | N | N | N | *Muscarinic cholinergic receptors belong to a larger family of G protein-coupled receptors* | Associated with psychiatric disease, alcohol use disorder and cognitive function (GWAS catalog) |  |
| rs12543318 | 8 | 88868340 | DCAF4L2 | Y | Y | Y | *DCAF4L2=Protein that regulates epithelial-mesenchymal transition (EMT) by activating NFkB signaling.* |  | Ludwig et al, 2017<br>Yu et al 2017<br>Leslie et al 2017<br>Ludwig et al 2012<br>Curtis et al 2021a)<br>Yang et al 2020<br>Mukhopadhyay et al, 2021 |
| rs17242358 | 8 | 129964873 | CCDC26<br>LINC00976 | Y | Y | Y | *Long Non-Protein Coding RNA* |  | Leslie et al, 2017<br>Curtis et al 2021 b)<br>Yu et al 2017<br>Grant et al 2009<br>Birnbaum et al 2009<br>Ludwig et al 2012 |
|  |  |  |  |  |  |  |  |  | Beaty et al 2010<br>Curtis et al 2021 a)<br>and b)<br>Awotoye et al 2022 |
| rs7870795 | 9 | 100574120 | FOXE1<br>PCTSC2 | N | Y | Y | *A thyroid transcription factor that plays a role in thyroid morphogenesis* | Involved in cleft palate formation. Nearest GWAS SNP rs12347191 in 9:100619719 (also sign in our analysis) | Castanet et al, 2002<br>Leslie et al, 2017<br>Yang et al 2020<br>Ray et al 2021 |
| rs1898349 | 10 | 118817067 | SHTN1 | Y | Y | Y | *Involved in positive regulation of neuron migration.* |  | Leslie et al, 2017<br>Huang et al 2019<br>Mangold et al 2010<br>Ludwig et al 2012<br>Curtis et al 2021 a)<br>and b)<br>Yu et al 2017<br>Sun et al 2015 |
| rs9601323 | 13 | 80700397 | SPRY2 | N | Y | Y | *Inhibitory activity on receptor tyrosine kinase signaling proteins. Bimodal regulator of epidermal growth factor receptor/mitogen-activated protein kinase signaling.* | Within credible region identified by Ludwig et al 2017, for SNP rs8001641 (also sign in our analysis) . Sole locus to have previously been identified with an effect on CLP but not CLO. | Ludwig et al, 2017<br>Yu et al 2017<br>Mangold 2010<br>Ludwig et 2012<br>Curtis et al 2021 a)<br>and b) |
| rs10483604 | 14 | 51485195 | TRIM9 | N | N | Y | *Member of the tripartite motif (TRIM) family, function not identified.* | Within 40kb of hit below | Ludwig et al, 2010<br>Ludwig et al, 2017 |
| rs4901118 | 14 | 51856109 | FRMD6 | Y | Y | Y | *FRMD6 =may be involved in maintaining cytoskeleton.* |  | Ludwig et al, 2017 |
| rs2600519 | 15 | 33063809 | FMN1 | N | Y | Y | *Protein has a role in the formation of adherens junction and the polymerization of linear actin cable.* | Associated with Robinow Syndrome, Autosomal Recessive 2 (RRS2) characteristics include facial dysmorphology (genecards) Within credible interval identified by Ludwig et al, for lead SNP rs1258763 at 33050423 (lead SNP in Ludwig et al, 2017 is also sign in our analysis) | Ludwig et al, 2017<br>Curtis et al 2021 a)<br>and b)<br>Mangold et al 2010<br>Ludwig et al 2012 |
| rs8044196 | 16 | 3984034 | CREBBP<br>ADCY9 | N | N | Y | *Transcription factor regulator, plays a critical role in embryonic development.* | Mutation in CREBBP recently identified in case with CPO. Within credible interval identified by Ludwig et al, for SNP rs8049367 at 3980445 (also sign in our analysis) | Wilson et al, 2023<br>Ludwig et al, 2017<br>Sun et al, 2015 |
| rs12971753 | 19 | 7499238 | ARHGEF1<br>8 | N | N | N | *Guanine nucleotide exchange factor (GEF) for RhoA GTPase. Regulation of actin cytoskeleton.* |  |  |
| rs2225351 | 20 | 6484640 | CASC20 | N | N | N | *Cancer susceptibility gene 20* | Region associated with facial morphology | *White et al 2020* |
| rs34753522 | 20 | 39278391 | MAFB | N | Y | Y | *MAF bZIP transcription factor B* | *MAFB* is a transcription factor | Leslie et al, 2017<br>Huang et al, 2019<br>Sun et al, 2015 |
| rs6018099 | 20 | 45473014 | EYA2 | N | N | Y | *Eyes absent 2 gene. Transcription factor involved in early embryonic development* | Expressed in early embryonic development. Region associated with cleft endophenotype | Indencleef et al, 2021,<br>Welzenbach et al, 2021 |

N= no previous evidence from GWAS, Y= previous evidence from GWAS

Lead SNPs rs4112328 (*LHX8)*, rs28361060 (*TSBP1*) and rs7870795 *(FOXE1*) had the smallest p-values in the “all cases” GWAS and were nominally significant (p<0.05) in the same direction across all subgroups. Most of the other SNPs we identified were genome wide significantly associated with cleft lip phenotypes or cleft palate only phenotypes but were not strongly associated with both and so the smallest p-values for these SNPs were not in “all cases”. After clumping, we identified 18 regions (2 mapping to *ARHGEF18* and *ARHGEF19* were novel) which showed the smallest p-values in the cleft lip with or without cleft palate (CL/P) or nsCL/P subtype GWAS and which were associated with both CLO and CLP. Additionally, rs62390705 (a novel region mapping to *FBN2*) was associated with CLO but not CLP (Cochrane’s Q p-value for heterogeneity = 0.0015), rs1347188 (*LINC01091*) was associated with CLP but not CLO (Cochrane’s Q p-value for heterogeneity =0.019) and finally rs9601323 (*SPRY2*) showed strong evidence of association with CLP (OR=1.46, p=1.01x10^-12^) but much weaker evidence of an effect on CLO (OR=1.14, p=0.04) (Cochrane’s Q p-value for heterogeneity =0.0027). All SNPs which were genome-wide significant for CL/P were also strongly associated (at least p<1x10^-6^) with nsCL/P, and vice versa.

Finally, we identified three novel loci primarily associated with cleft palate only (CPO). Lead SNP rs72823645 (*SLC35B3)* was associated with both non-syndromic cleft palate only (nsCPO) and Pierre Robin sequence (PRS) (albeit not genome wide significant in PRS, p=2.82x10^-5^), as well as being nominally associated with cleft lip phenotypes with attenuated effects in the same direction. Lead SNP rs17168736 (*CHRM2)*, was strongly associated with nsCPO and weakly with cleft lip phenotypes in the same direction but was not associated with PRS. The third lead SNP, rs2225351 (*CASC20)* was genome wide significant for CPO and PRS, with a weaker effect on nsCPO, and no association with cleft lip phenotypes. Table 3 shows a summary of the evidence linking each of the GWAS significant regions to orofacial cleft. Table 3 shows a summary of the evidence linking our lead SNPs to orofacial clefts.

### Pierre Robin Sequence (PRS)

Although we identified only one genome-wide significant region for PRS (*CASC20*), we found 21 additional genomic regions with suggestive associations in our case-control analysis (p<1x10^-5^) (Table 4). The identified regions have not previously been linked to orofacial clefts in GWAS. Several of these regions or mapped genes however have been associated with facial morphology in GWAS (Li et al., 2023, Hoskens et al., 2021, Indencleef et al., 2021), linked to syndromes characterised by facial features (Kent et al., 2020), or have shown evidence of involvement in orofacial clefts in animal studies (Fairchild and Gammill., 2013, Sandford et al., 1997).

**Table 4.**
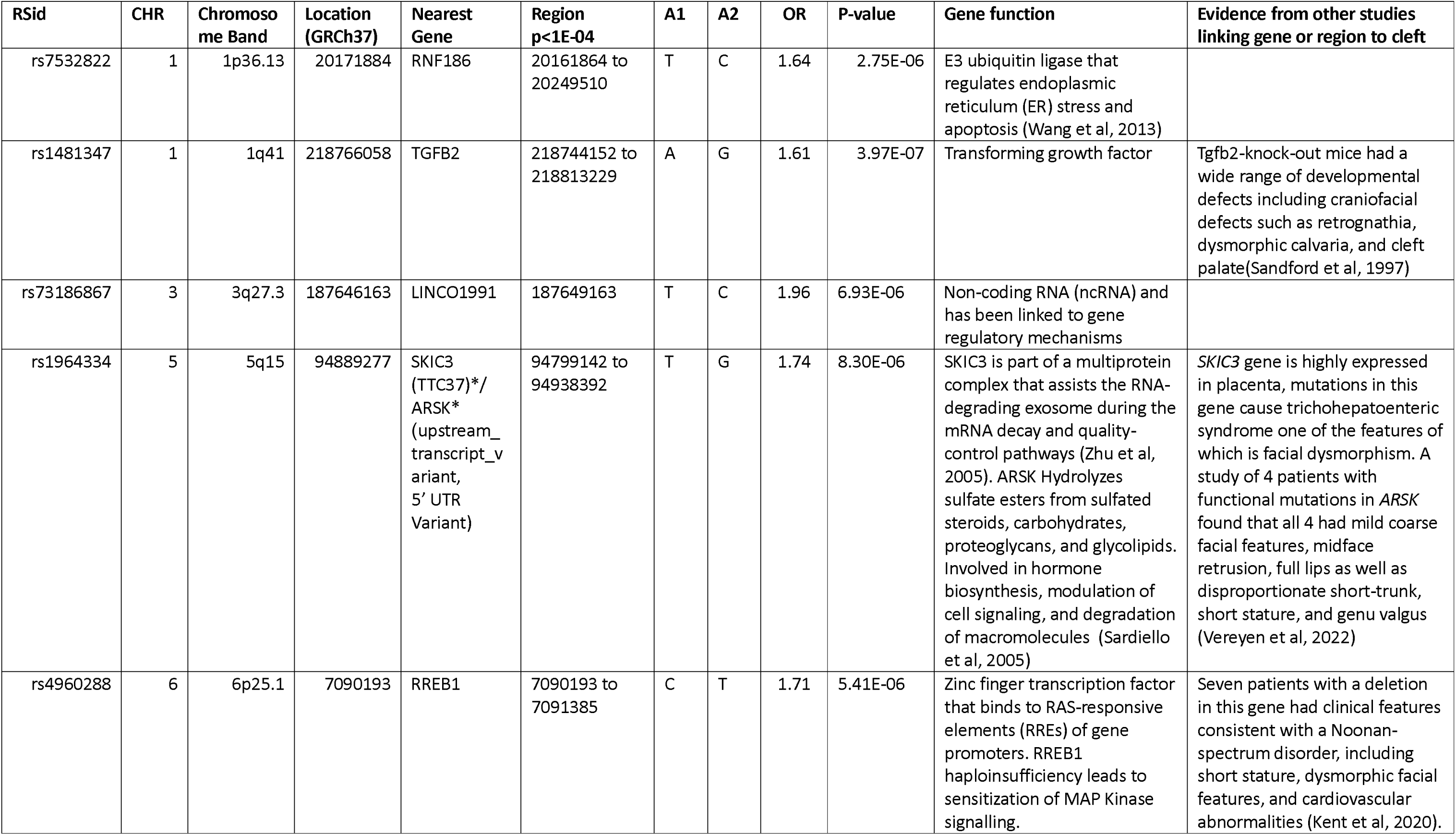
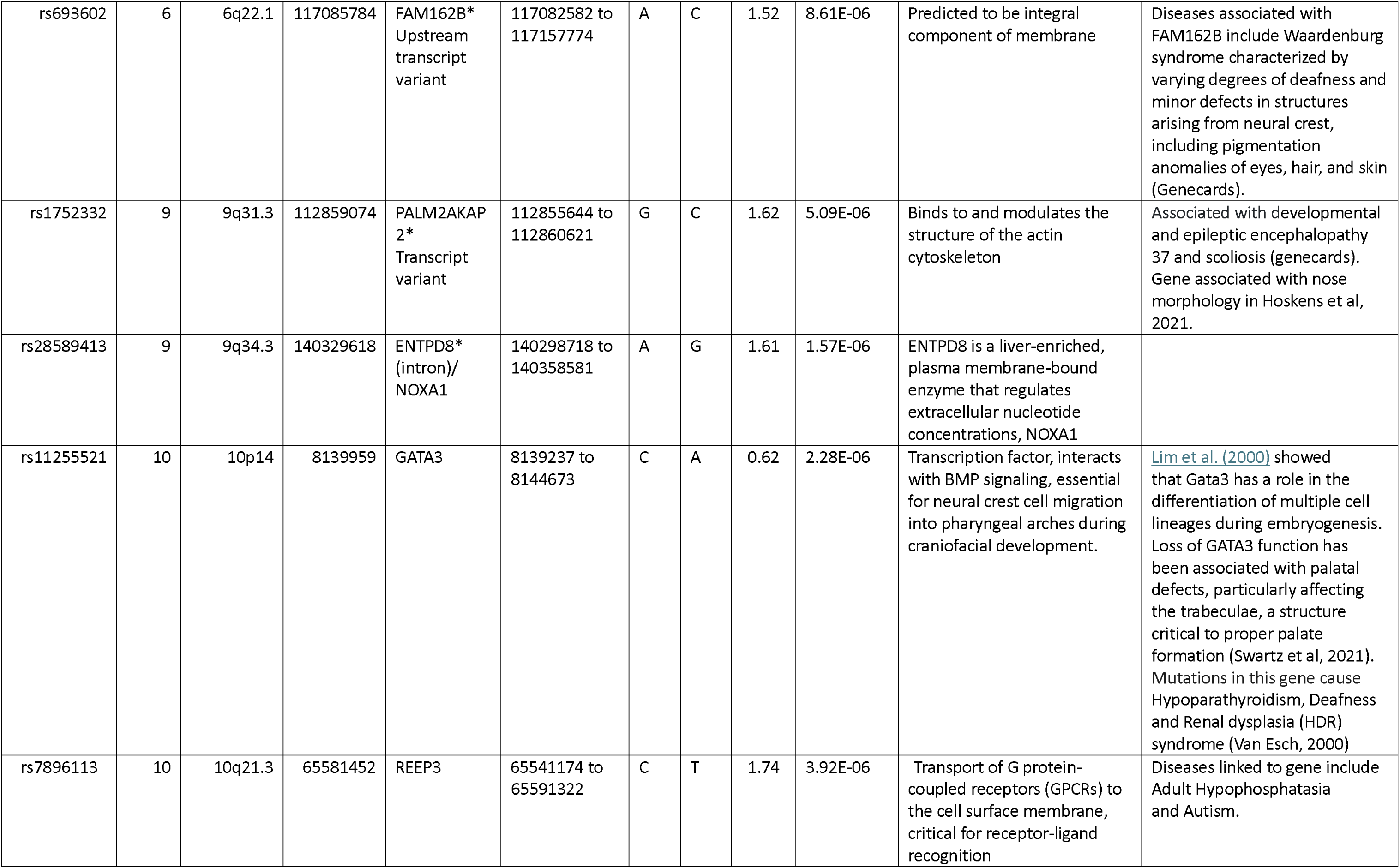
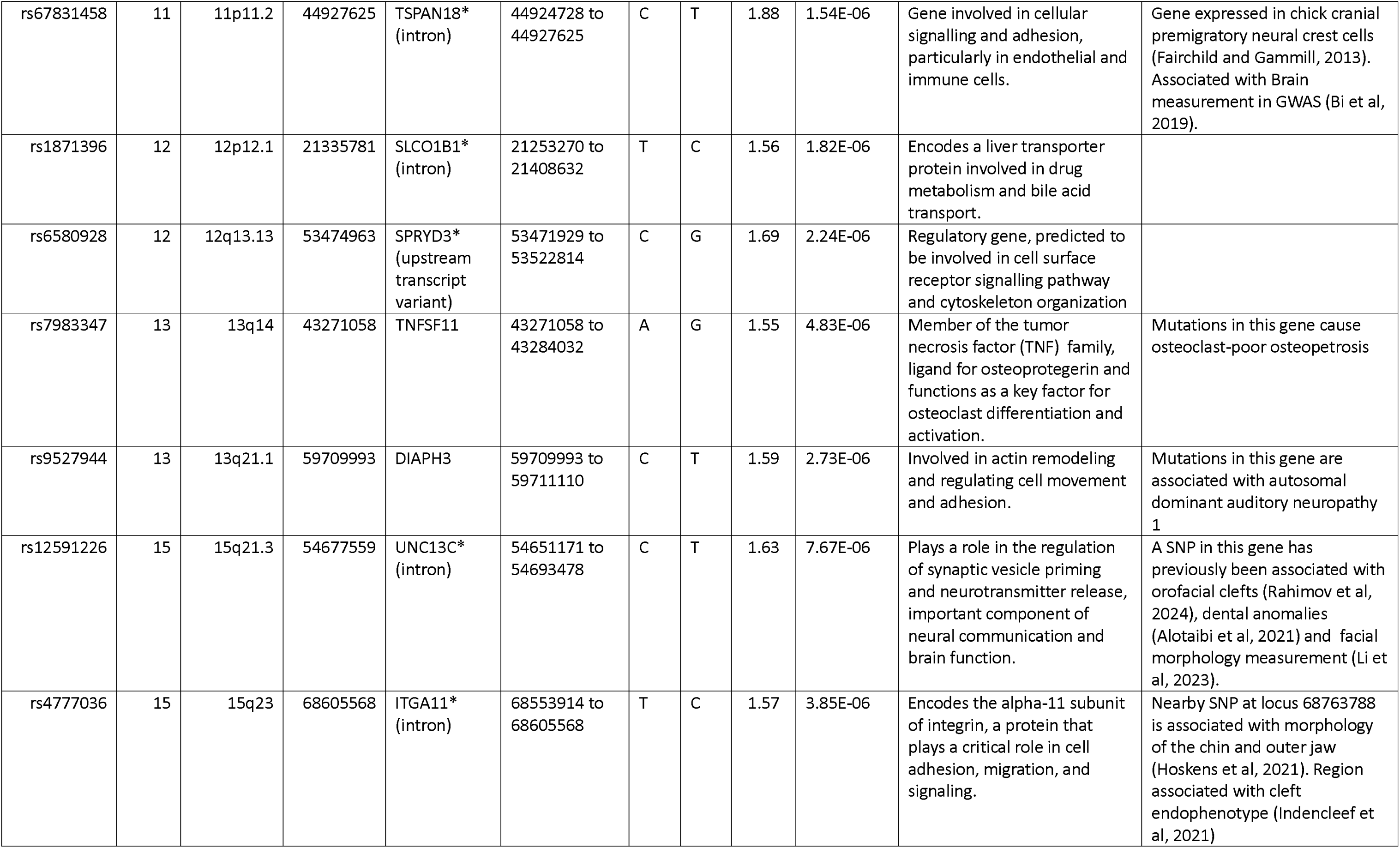
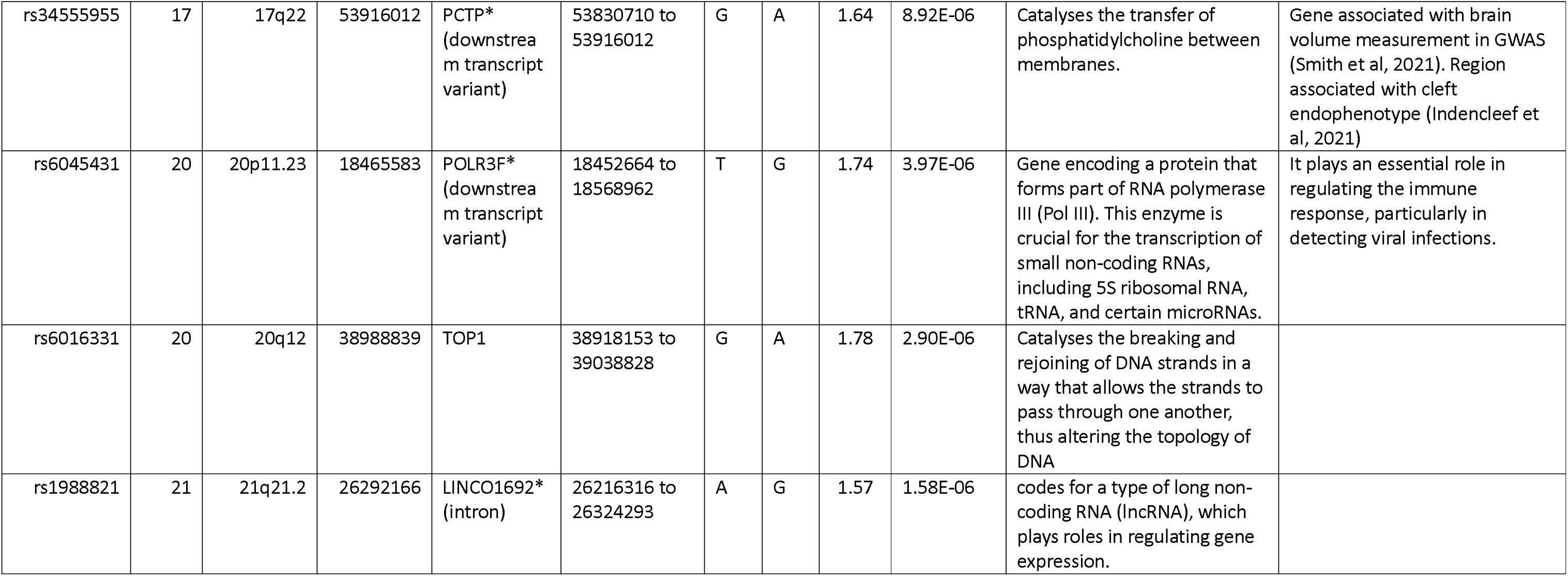
Results of clumped SNPs showing suggestive associations (p<1x10^-5^) with Pierre Robin Sequence in case-control analysis.

**Table 5.**
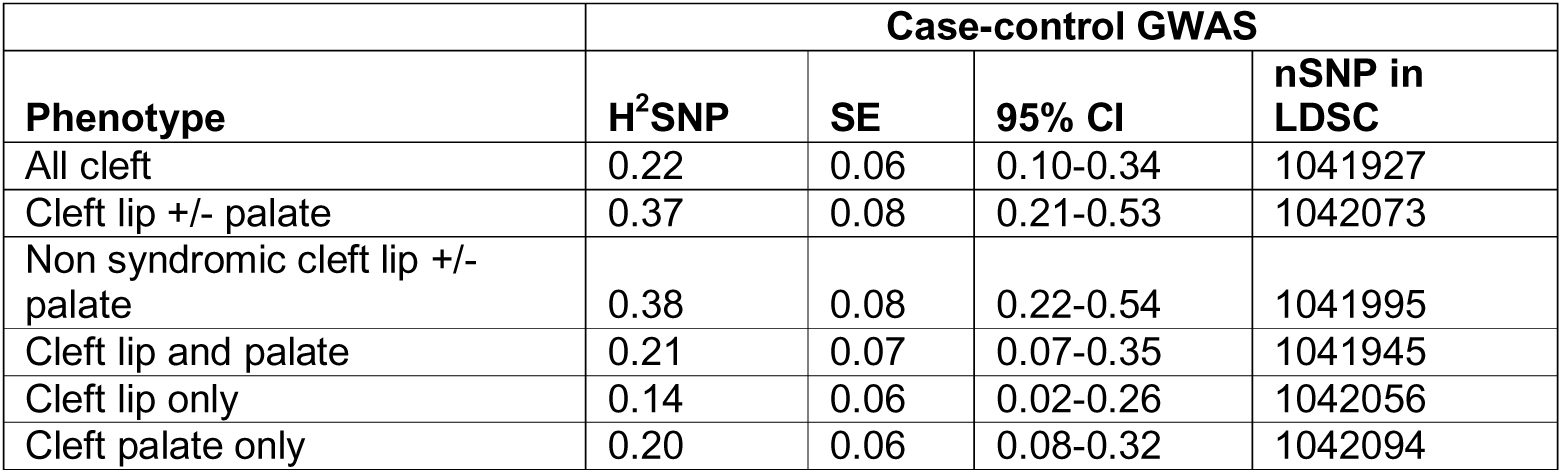
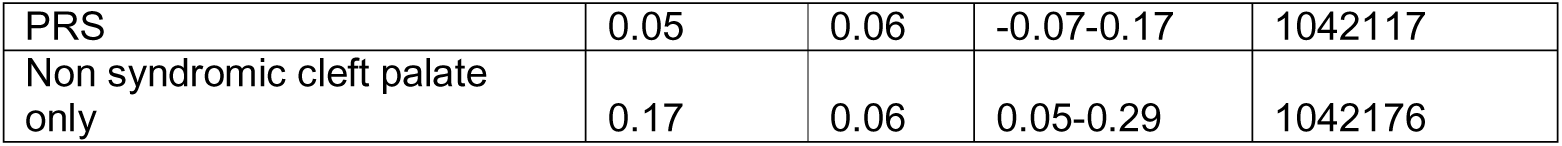
SNP heritability estimated using LD score regression.

Supplementary Figures 2-17 show Manhattan and QQ plots of our analyses.

### Replication

Almost all of our lead SNPs with the strongest evidence of association in CL/P or nsCL/P were replicated (with at least nominal levels of significance p<0.05 and effects in the same direction) in the meta-analysis by Welzenbach et al. (2021) (see Table 2), with some also being GWAS-significant in that study. Two novel SNPs which were genome wide significant in nsCL/P (rs61769781) and CLO (rs62390705) showed some evidence of association with nsCL/P in the replication meta-analysis. Although, rs12971753 (*ARHGEF18*) did not replicate, this SNP showed weak evidence in the same direction. Of the three SNPs with the strongest evidence in the “all clefts” group, two were not associated with nsCL/P in the replication sample; however, rs7870795 (*FOXE1*) showed strong evidence of association. As expected, our CPO hits were not associated with nsCL/P in the replication sample. We wanted to replicate our CPO hits in CPO GWAS, however no GWAS of all CPO (syndromic and non-syndromic cases) or of PRS were available for this purpose and our nsCPO hit was not available in a recent meta-analysis of nsCPO (Jia et al, 2024).

### Correlation across subgroups

Figure 2 shows a forest plot of effect sizes and confidence intervals for each of our 27 genome wide significant hits across all subgroups. For the most part, SNPs primarily associated with cleft lip phenotypes were not associated with cleft palate only phenotypes. However, three SNPs showed some weak evidence of an effect on PRS in the opposite direction to their effect on cleft lip phenotypes: rs62390705 (p = 0.05), rs126280 (p = 0.002), and rs4952552 (p = 0.04). Along with the three SNPs which showed the strongest evidence of effect across all clefts, there were another four SNPs (rs1347188, rs17168736, rs2600519, rs72823645) which showed some evidence of association in the same direction across all phenotypes.

**Figure 2.**
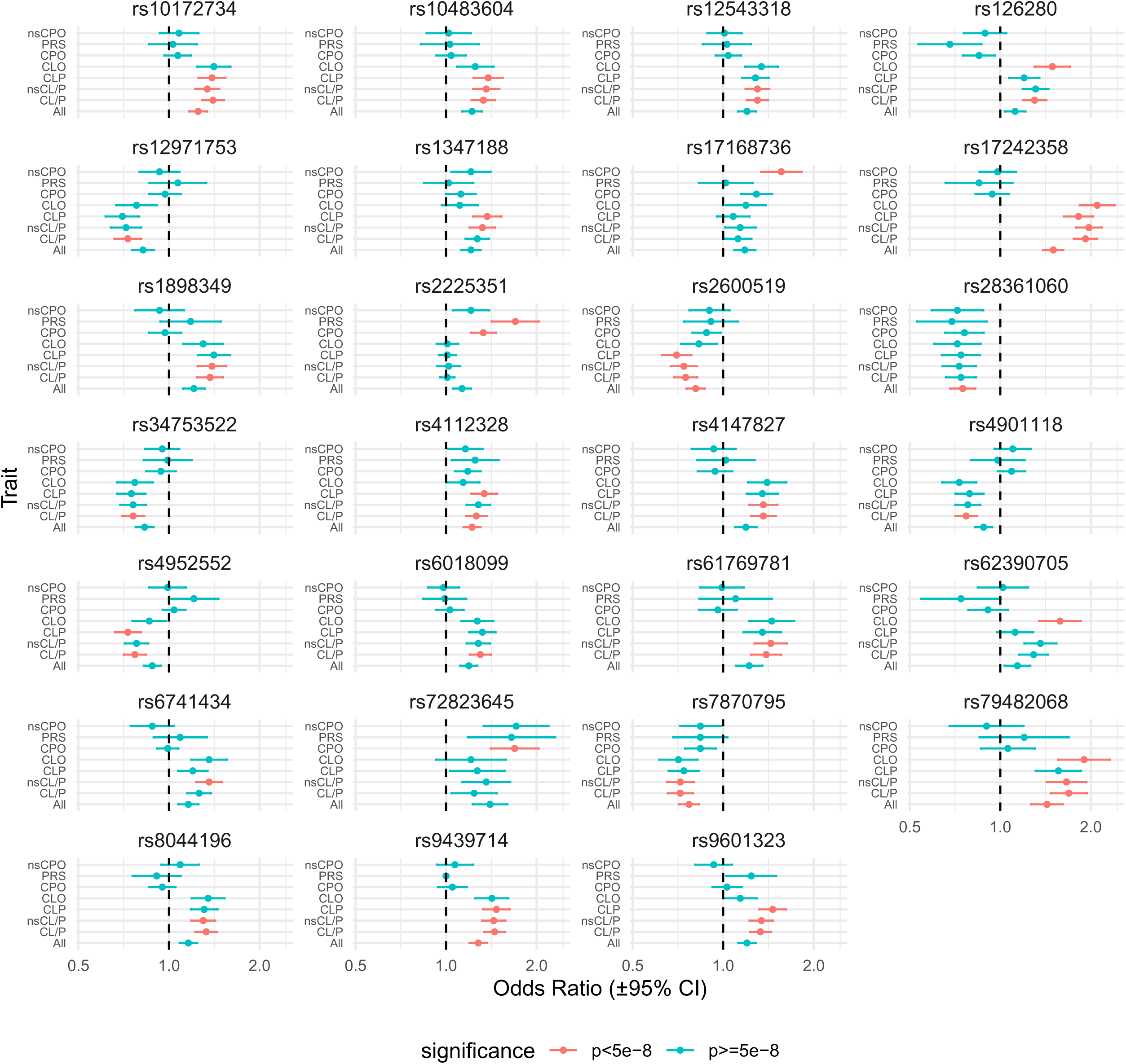
Forest plot of our showing SNP effects across all subgroups for the 27 SNPs reaching genome wide significance in at least one of our GWAS

Our correlation matrix of effect estimates for lead SNPs across subgroups (Figure 3) revealed very strong correlations (> 0.86) between the various cleft lip phenotypes. Despite CLO and CLP being mutually exclusive, they still showed a high correlation. In contrast, correlations between cleft palate only and cleft lip subgroups were much weaker, with a weak negative correlation between CLO and PRS (Pearson’s correlation coefficients ranged from -0.03 to 0.24). Within the CPO subgroups, the correlation between nsCPO and PRS was higher (0.57) than the correlation between CPO and cleft lip subtypes, but indicated that these phenotypes are likely to have different underlying genetic causes. For CL/P, there was almost perfect correlation between all CL/P cases and a subgroup which excluded syndromic cases- nsCL/P (correlation coefficient = 0.99). Whilst there was also a strong correlation between all CPO cases and the subgroup excluding syndromic cases (correlation coefficient =0.81) there was evidence of some differences between the two.

**Figure 3.**
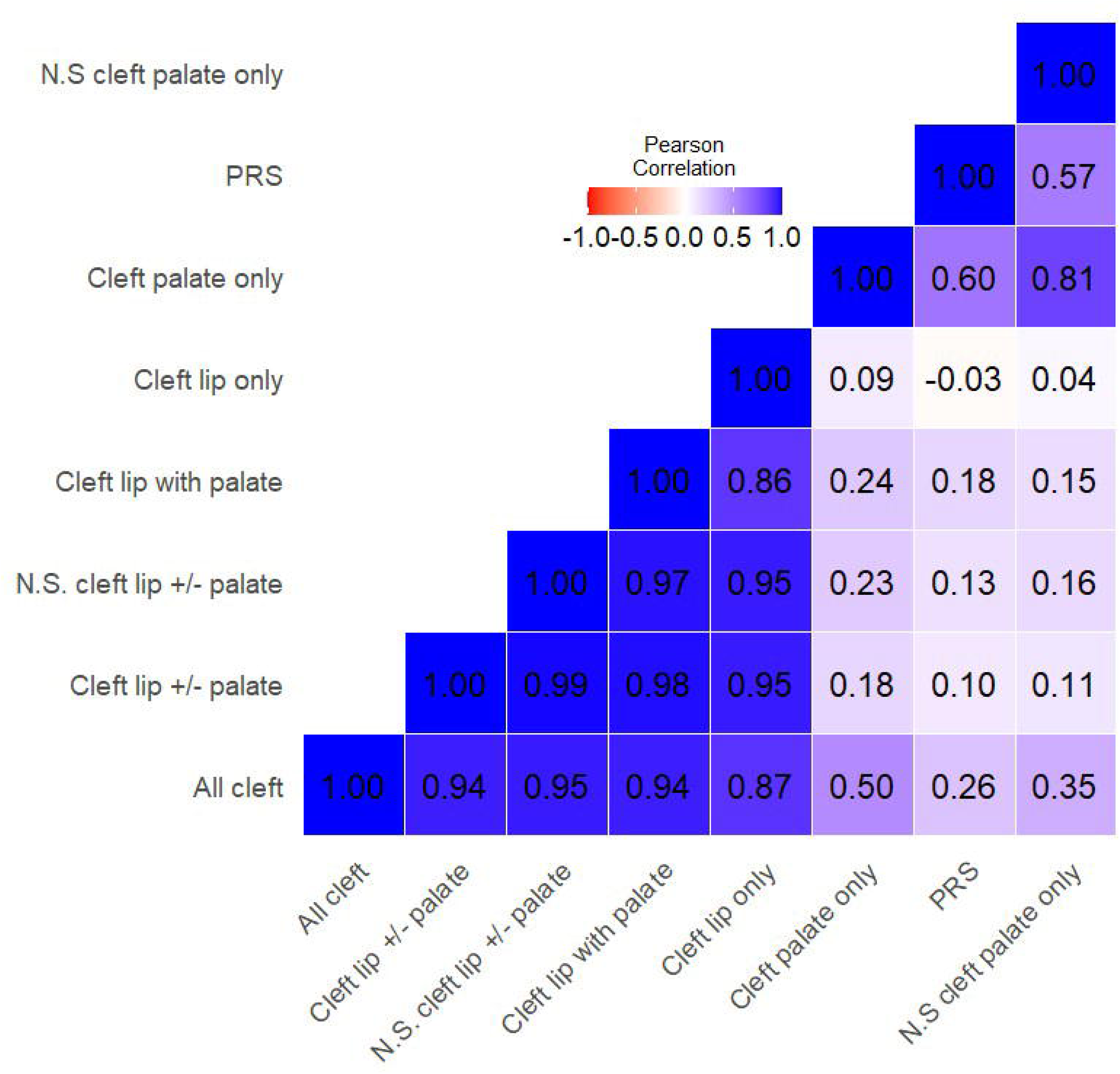
Heatmap showing correlation coefficients based on effect sizes of our 27 lead genome wide significant SNPs across subtypes

### Proportion of variance explained

Our estimates for SNP heritability (calculated using LD-score regression) based on our case-control analysis ranged from 5% (95%CI: -0.07 to 0.17) for PRS to 38% (95%CI: 0.22 to 0.54) for nsCL/P, with the SNP heritability of nsCPO estimated to be around 17% (95%CI: 0.05 to 0.29) (Table 3).

### Enriched biological processes

The biological processes which were most enriched in the gene set which mapped to all 27 genome wide significant hits from our case-control analyses were limp morphogenesis (5 genes 25.07 fold enrichment, p=1.46x10^-6^, false discovery rate (FDR) adjusted p=0.007) and embryo development (10 genes, 7.29 fold enrichment, p=4.17x10^-7^, FDR adjusted p=0.006). Within the gene set identified by the PRS GWAS none of the biological processes showed evidence of enrichment.

## Discussion

Our study represents a comprehensive set of GWAS of orofacial clefting, in which we performed GWAS of all clefts combined plus GWAS of 7 orofacial cleft subgroups. We identified several novel loci, replicated key associations from previous studies, and provided new insights into the genetic architecture of orofacial clefts.

### Key findings, novel associations and consistency with other evidence

We identified 27 genome-wide-significant loci, 8 of which were novel. Novel associations were identified between SNPs mapping to *ARHGEF18* and just upstream of *ARHGEF19* and CL/P.These genes are involved in Rho GTPase signalling pathways, which regulate neural crest cell migration and cytoskeletal organization—processes critical for craniofacial structure development and fusion (Blomquist et al., 2000; Gossen et al., 2024). We have previously shown *ARHGEF19* to be associated with facial morphology (right endocanthion to left cheilion distance) (Xiong et al., 2021).

*CASC20* has been previously linked to cleft lip, but we did not find an association between SNPs at this locus and cleft lip and to our knowledge ours was the first study to associate it with cleft palate, and specifically to PRS. *CASC* genes are classified as cancer susceptibility genes and may therefore influence cell growth and proliferation, although their functional roles are currently unclear.

Prior GWAS have not typically been carried out on all orofacial cleft cases but instead have tended to be restricted to non-syndromic cleft lip with or without palate or cleft palate cases only. To our knowledge there is only one GWAS study published in 2017 which combined cleft lip and cleft palate only subtypes, although this study did not include syndromic cases (Leslie et al, 2017). Our study suggests that GWAS of all cases combined may also be fruitful and future studies, which include all orofacial clefts, with larger sample sizes than ours may find more variants. Three genomic regions mapping to *LHX8, FOXE1*, and *TSBP1* showed the strongest evidence of association when all clefts were combined. All three regions are transcription and may therefore have wide ranging effects on development. *LHX8* and *FOXE1* are transcription factors essential for craniofacial development and have been linked to cleft palate (Zhao et al., 1999; Meng et al., 2012). Whilst SNPs in the *FOXE1* region have been frequently associated with different subgroups of orofacial clefts in GWAS and SNPs in this region were genome wide significantly associated with all orofacial clefts in the GWAS by Leslie et al. (e.g., Leslie et al., 2017; Yang et al., 2020, Ludwig et al., 2014), *LHX8* has not been previously been identified in orofacial cleft GWAS. However, a sequencing study did detect a mutation in *LHX8* in a CLP patient (Vieira et al., 2005). *TSBP1* is less well-studied but is thought to regulate transcription, and its promoter region includes binding sites for forkhead (FOX) proteins (Fishilevich et al., 2017). Additionally, *TSBP1* is located upstream of *NOTCH4*, a gene implicated in craniofacial development (Pakvasa et al., 2021). Two further previously identified regions and two novel regions were also nominally associated with all cleft types in the same direction (*LINCO1091, FMN1, SLC35B3, PSMCIP3*), albeit for these SNPs they exhibited genome wide significant associations with either lip or palate only phenotypes, but were only nominally significantly associated with the other phenotype. *SLC25B3*, was a novel locus associated with CPO in our GWAS (and to a lesser extent with nsCL/P), it is a solute transporter gene linked to disorders featuring cleft palate and facial dysmorphism (Paganini et al., 2020). Similarly SNPs mapping to *PSMCIP3* were associated with nsCPO and also nominally associated with CLO in the same direction.

SNPs upstream of the *FBN2* gene were associated with CLO in our analysis. *FBN2* encodes a component of connective tissue microfibrils and has been associated with lip morphology (White et al., 2020; Hoskins et al., 2021), although our study is the first to associate *FBN2* with orofacial clefts. We found two regions to be genome wide significant in our CLP subgroup, but to only show very weak evidence of association with CLO, one of these regions maps to *SPRY2*, the CLP-specific effect of this locus has been shown in previous analyses (Ludwig et al. 2012, Jia et al. 2015).

### PRS findings

To our knowledge this was the first GWAS of PRS to date. Our sample size for this subgroup was small, just 237 cases and we were therefore underpowered to detect many potential associations. Nonetheless we did identify one GWAS significant SNP mapping to *CASC20*. We also found a further 21 regions where associations approached GWAS significance. Only one of these regions (UNC13C) had previously been identified in a cleft GWAS, with two more genes associated with facial morphology (including a cleft endophenotype) in GWAS (*ITGA11, UNC13C* and *PCTP*) (Li et al., 2023, Hoskens et al., Indencleef et al., 2021). In addition, *GATA3* and *TGFB2* have been shown to be involved in palate formation (Van Esch et al., 2000 and Sandford et al., 2017). A further five regions mapped to genes associated with syndromes which have craniofacial anomalies as a feature (*TTC37, RREB1, FAM162B, PALM2AKAP and NOXA1*) (da Silva et al., 2025, Kent et al., 2020, www.genecards.org). These genes warrant future research, although replication in larger GWAS is required.

### Phenotypic and Genetic Correlations

The correlation matrix of effect estimates across subgroups revealed strong genetic overlap within cleft lip phenotypes, consistent with shared genetic underpinnings. The weaker correlations between cleft lip and cleft palate phenotypes support their classification as distinct entities. Notably, some SNPs exhibited opposite effects on PRS compared to cleft lip phenotypes, suggesting that the two subtypes might be two opposite extremes resulting from disruptions to shared developmental processes.

Incorporating syndromic cases into our CL/P GWAS analysis had minimal impact on SNP effect estimates, as indicated by a high correlation (r = 0.99) between the full dataset and the subset excluding syndromic cases, whereas including syndromic cases increased the sample size by 26%. These results suggest that (a) restricting the analysis to non-syndromic cases is unnecessary and (b) common genetic variants may contribute similarly to both syndromic and non-syndromic cases. Our findings align with Wilson et al. (2021), who reported that the distinction between these groups is not well-defined. Given that information on syndrome diagnosis is sometimes not available and potentially problematic, and given that syndromes are often underdiagnosed (Davies et al., 2024b), it is reassuring to know this is unlikely to influence results.

Our SNP heritability estimates found that 38% of the variation in nsCL/P and 17% of the variation in nsCPO was explained by common genetic variation. Our SNP heritability estimate for nsCL/P was slightly higher than we estimated in 2017 (32% (standard error = 8.5%)) based on another cohort (Welzenbach et al, 2021), although the confidence intervals overlap. To our knowledge SNP heritability for nsCPO has not previously been estimated.

### Strengths and limitations of the study

Our study has several notable strengths. First, we utilized a well-characterized case population with detailed classification of cleft subtypes. Combined with a large, population-based control group at an approximate 4:1 control-to-case ratio, this significantly enhanced the statistical power of our analysis.

Another strength of our study was the ability to compare genetic effect estimates across multiple cleft subtypes. This analysis revealed risk loci shared across all cleft subtypes as well as those specific to cleft lip only, cleft palate only, or cleft lip and palate. Furthermore, the cross-subtype comparison identified genes exhibiting opposing effects on PRS and cleft lip. This intriguing finding warrants further exploration to identify the mechanism involved.

Our study had several limitations, the most significant being the small number of cases in many subgroup analyses, particularly for PRS. To address this, we focused not only on p-values but also on effect sizes across subgroups for SNPs that reached genome wide significance in at least one analysis and included a replication sample. Despite these efforts, we likely missed many regions associated with cleft.

We investigated the correlation between subtypes with and without syndromes, however we expected these to be correlated to a certain extent as one is a subset of the other. Ideally we would have investigated the correlation of non-syndromic and syndromic forms of CL/P, but we didn’t do this because the sample size for syndromic cases with a cleft lip was small. In addition, we were unable to use whole genome methods to estimate genetic correlation and some of the correlation coefficients between subgroups are likely to have been affected due to random fluctuations in effect estimates, both due to small sample size. In the future, we plan to enhance our analyses by integrating data from other cleft studies through meta-analyses to increase sample sizes.

Many of the SNPs we identified were located in non-coding regions of the genome and are unlikely to be the causal variants. In addition, we mapped SNPs to the nearest gene, but this is not necessarily the gene involved in orofacial clefts. For example SNPs, such as the ones we identified, in the *ABCA4* gene have previously been shown to be in enhancer regions which control *ARHGAP29* expression and evidence suggests that it is *ARHGAP29* not *ABCA4* that plays a role in orofacial clefting (Lui et al, 2017). Future studies are required to pinpoint causal variants in these regions and to clarify the biological processes through which the associated genes influence cleft formation.

## Conclusion

The identification of novel loci and the replication of known regions highlights the robustness of our approach. The novel associations, particularly for CPO and PRS expand the genetic landscape of orofacial clefts and may inform future studies of craniofacial development. Functional validation of the implicated genes and loci, particularly those with pleiotropic effects, is a critical next step.

Larger studies with more diverse populations which follow our approach are likely to capture even more genetic variation contributing to orofacial clefts. The interplay of genetic and environmental factors, particularly for rarer phenotypes such as PRS remains an important area for future research.

## Materials and Methods

### Ethical approval

All study participants were recruited with informed parental consent. Ethical approval was granted for the Cleft Collective by the NRES committee South-West (13/SW/0064). The Millennium cohort study obtained ethical approval from London-Central Research Ethics Committee (13/LO/1786).

### Study population and sample

#### Orofacial cleft families

Children born with an orofacial cleft and their families who were registered at one of 17 cleft centres across the UK were recruited to the Cleft Collective Cohort study between November 2013 and June 2020. Parents who consented to participate were sent a questionnaire to complete and were asked to provide a saliva sample, both of which were returned by post, the questionnaires to the Cleft Collective team and the biological samples to the Bristol Bioscience Laboratories . For children recruited before their first surgery, blood and tissue samples were collected during the procedure. For those cases recruited at age 5, saliva samples were obtained at the five-year audit clinic.

#### Controls

Controls were from the Millenium Cohort Study, a population-based study of children who were born across the UK over a 17 month period (September 2000-January 2002). The study is coordinated from the University College London (Connelly and Platt, 2014), although processing of the biological samples was performed at the University of Bristol. Saliva samples were collected from 9259 children in the cohort during a home visit by the study team in 2015-2016 when they were approximately 14 years old (Fitzsimons et al., 2022).

### Case definition

Information on cleft subtypes including syndrome status came from either the parent questionnaire, forms completed by surgeons at the time of cleft surgery or from clinical notes. We initially grouped all cases together to form a group labelled “all clefts”. In addition, we performed orofacial cleft subtype analyses of seven different subtypes as detailed in Table 1.

All biological samples from the Cleft Collective and the Millenium Cohort Study were sent to Bristol Bioresource Laboratories (https://www.bristol.ac.uk/population-health-sciences/research/groups/bblabs/) where they were stored and processed, and where genotyping was carried out (Fitzsimons et al., 2022, Davies et al., 2024).

For information on DNA extraction and quantification, please see **Supplementary methods**.

### Genotyping

Genotyping was performed on 200ng of DNA using the Illumina Infinium global screening array-24 v1.0 in the Millennium cohort (606,434 SNPs) and using the Illumina Infinium global screening array-24 v3.0 in the Cleft Collective (645,027 SNPs). Different arrays were used for the cases and controls because genotyping was performed at a later date for the Cleft Collective. Quality control (QC) steps were performed and samples were excluded if they had a within sample SNP call rate <97%, biological sex mismatch (between records and DNA) and low child-parent inter-relatedness based on identity by descent (IBD<0.4). SNPs were excluded based on call rate <98%, minor allele frequency (MAF) <1%, Hardy-Weinberg equilibrium P-value < 1x10^-6^ in controls or poor clustering and excess heterozygosity in cases (excess het > +/-0.3). After QC 457,642 SNPs genotyped in controls and 480,916 SNPs genotyped in cases remained. Figure 1 shows the number of individuals excluded at each stage of the QC process and the number of included cases for each orofacial cleft subtype.

#### Merged cases and controls

Cases and controls were merged and 399,882 SNPs common to both groups were retained. Principal components (PC) for ancestry were generated and applied to the combined group, and outlier filtering applied. Supplementary Figure 1 shows the distribution of the included cases and controls in a scatter plot of PC1 versus PC2.

### Imputation

Imputation was performed on the merged case-control dataset using the TopMed Imputation Server (Taliun et al, 2021) to the TopMed R3 reference panel of 133,597 samples. The imputation used Minimac4, and phasing with Eagle v2.4. Following imputation, SNPs were excluded if imputation quality RSq score < 0.3, MAF < 0.01, or they were monomorphic.

Following imputation, SNPs with call rate < 98% were identified and excluded (n = 166), and additional filtering was performed on SNPs with HWE P < 1x10^-6^ in controls (n = 399,457), leaving 7,606,581 SNPs which passed our quality control steps. After excluding SNPs with a minor allele frequency of <0.05, a maximum of 5,457,364 SNPs were included in our analyses.

### Statistical analysis

Genome-wide association analyses were conducted comparing cases with controls. Initially, all cleft subtypes were combined to maximize statistical power. Subsequently, GWAS were performed for individual cleft subtypes as defined above. Whilst we used a common set of controls across all our analyses the number of cases varied in the different analyses (see Figure 1 for number of cases in each subtype). Finally, we explored the extent to which genetic variants are shared between the subtypes and the heterogeneity between them. PLINK v1.9 (Purcell et al., 2007) was used to perform genome-wide association tests.

Allele frequency differences between cases and controls were tested using logistic regression adjusted for scores of the first 10 principal components generated from our merged case-control pre-imputed data. SNPs were included if MAF > 0.05 in the specific phenotype subset. This analysis was performed using data imputed using the TopMed R3 reference panel (McCarthy et al, 2016, Taliun et al., 2021).

#### Subtype comparisons

Results for any SNP that was genome-wide significant (5 x10^-8^) in at least one subgroup were reported across all subgroups. We did not make any adjustment for multiple testing due to the inclusion of multiple subgroups because the subgroups were not independent of each other. However, we examined the differences in p-values and effect estimates between subgroups, and plotted these as a forest plot as part of our analysis. We also reported the results for the orofacial cleft subgroup with the lowest p-value for each SNP plus any other subgroups with p-values of <1 x10^-4^ in Table 2. We relaxed our threshold for reporting associations with secondary phenotypes because some subgroups had smaller numbers and hence lower power to detect similar effects. We conducted pairwise comparisons across subtypes by calculating Pearson’s correlation coefficients using effect estimates for any clumped SNP that was GWAS significant for at least one cleft subtype. We also performed a meta-analysis and used a Cochrane’s Q-test to test whether there was evidence for heterogeneity between cleft lip only (CLO) and cleft lip and palate (CLP) subtypes for the following SNPs: rs1347188, rs62390705, rs9601323 which appeared to show a difference in associations between the two subtypes.

#### Replication

A look-up was performed by extracting the results for the lead SNPs for each genome-wide significant locus in our analyses from a recent independent meta-analysis of non-syndromic cleft lip with or without cleft palate (nsCL/P) (Welzenbach et al, 2021) and we reported the results alongside our results. The meta-analysis combined case-control and trio data from 3 separate studies of nsCL/P (Mangold et al, 2010; Beaty et al, 2010 and Leslie et al, 2016) comprising of individuals of European, Asian and Latin American ancestry. The final dataset comprised 1,247 nsCL/P cases, 2,879 controls, 2,699 case-parent trios, and approximately 7.74 million SNPs. We reported the p-value of the association of the SNP with nsCL/P in our replication sample alongside our main results. Where the SNP effect was in the same direction and with a p-value <0.05 we considered this a positive replication.

#### Follow-up analyses

There were 8 sets of summary statistics generated by the GWAS of the 8 phenotypes. For each set of results, Manhattan and QQ plots were generated in R version 4.3.1 using the package *qqman* (Turner et al, 2018). QQ plots were used to assess inflation, along with lambda estimates. Genome-wide significant SNPs (P < 5x10^-8^) were identified, and genetically independent locations identified using the package *ieugwasr* (Hemani et al., 2024) to clump SNPs with a clumping window of 250kb and r2 cutoff of 0.2.

Summary statistics were uploaded to FUMA SNP2GENE (Watanabe *et al.,* 2017) for functional mapping and annotation, and LocusZoom (Pruim et al., 2010) to generate plots of locations with genome-wide significant associations. On visual inspection of the locus zoom plots, some clumped regions were merged or separated. The lead SNP in each region was reported as the one with the smallest p-value. The 95% credible region was also noted.

Mapped genes were either genes which contained the SNP or the gene which was closest to the SNP according to dbSNP (https://www.ncbi.nlm.nih.gov/projects/SNP/).

Clumped SNPs, their mapped genes and a region of 500,000 bases either side of the location of the SNP were searched in GWAS catalog (<u>GWAS Catalog</u>) and any associations with orofacial clefts were noted. We also used https://www.geneontology.org/ to investigate whether there were any biological processes which were over-represented among the mapped genes.

Finally, the measured SNP-based heritability was quantified for each analysis, by LD score regression (Lee et al, 2018). This involved regressing the SNP-level summary statistics for a phenotype against the standardised sum of LD.

## Data Availability

All data produced in the present study are available upon submitting a request to The Cleft Collective Project Management Group (https://www.bristol.ac.uk/dental/cleft-collective/professionals/access/)

## Acknowledgments

We are grateful to the families who participated in this study, the UK NHS cleft teams, and the Cleft Collective team, who helped facilitate this study. The views expressed in this publication are those of the authors and not necessarily those of the funders or the Cleft Collective Cohort Studies team. SJL received support for this study from an MRC project grant (MR/T002093/1). ES receives funding from an MRC project grant MR/W020297/1. Initial funding for the Cleft Collective was provided by the Scar Free Foundation who also provided separate funding to genotype study participants; additional funding was provided by The Underwood Trust and the Vocational Training Charitable Trust (VTCT). GDS is director of the Medical Research Council Integrative Epidemiology Unit at the University of Bristol which is supported by the Medical Research Council (MC_UU_00032/1)

## References

1. Alcaraz, W. A., Gold, D. A., Raponi, E., Gent, P. M., Concepcion, D., & Hamilton, B. A. (2006). Zfp423 controls proliferation and differentiation of neural precursors in cerebellar vermis formation. Proceedings of the National Academy of Sciences of the United States of America, 103(51), 19424–19429. 10.1073/pnas.0609184103

2. Alotaibi RN, Howe BJ, Moreno Uribe LM, Ramirez CV, Restrepo C, Deleyiannis FWB, Padilla C, Orioli IM, Buxó CJ, Hecht JT, Wehby GL, Neiswanger K, Murray JC, Shaffer JR, Weinberg SM, Marazita ML. Multivariate GWAS of Structural Dental Anomalies and Dental Caries in a Multi-Ethnic Cohort. Front Dent Med. 2021;2:771116. doi: 10.3389/fdmed.2021.771116.

3. Awotoye, W., Comnick, C., Pendleton, C., Zeng, E., Alade, A., Mossey, P. A., Gowans, L. J. J., Eshete, M. A., Adeyemo, W. L., Naicker, T., Adeleke, C., Busch, T., Li, M., Petrin, A., Olotu, J., Hassan, M., Pape, J., Miller, S. E., Donkor, P., … Butali, A. (2022). Genome-wide gene-by-sex interaction studies identify novel nonsyndromic orofacial clefts risk locus. Journal of Dental Research, 101(4), 465–472. 10.1177/00220345211046614

4. Beaty, T. H., Murray, J. C., Marazita, M. L., Munger, R. G., Ruczinski, I., Hetmanski, J. B., Liang, K. Y., Wu, T., Murray, T., Fallin, M. D., Redett, R. A., Raymond, G., Schwender, H., Jin, S. C., Cooper, M. E., Dunnwald, M., Mansilla, M. A., Leslie, E., Bullard, S., … Scott, A. F. (2010). A genome-wide association study of cleft lip with and without cleft palate identifies risk variants near MAFB and ABCA4. Nature Genetics, 42(6), 525–529. 10.1038/ng.580

5. Bi, X., Feng, L., Wang, S., Lin, Z., Li, T., Zhao, B., Zhu, H., & Zhang, H. (2019). Common genetic variants have associations with human cortical brain regions and risk of schizophrenia. Genetic Epidemiology, 43(5), 548–558. 10.1002/gepi.22203

6. Birnbaum, S., Ludwig, K. U., Reutter, H., Herms, S., Steffens, M., Rubini, M., Baluardo, C., Ferrian, M., Almeida de Assis, N., Alblas, M. A., Barth, S., Freudenberg, J., Lauster, C., Schmidt, G., Scheer, M., Braumann, B., Bergé, S. J., Reich, R. H., Schiefke, F., … Mangold, E. (2009). Key susceptibility locus for nonsyndromic cleft lip with or without cleft palate on chromosome 8q24. Nature Genetics, 41(4), 473–477. 10.1038/ng.333

7. Blomquist, A., Schwörer, G., Schablowski, H., Psoma, A., Lehnen, M., Jakobs, K. H., & Rümenapp, U. (2000). Identification and characterization of a novel Rho-specific guanine nucleotide exchange factor. The Biochemical journal, 352 *Pt* 2(Pt 2), 319–325.

8. Bonfante, B., Faux, P., Navarro, N., Mendoza-Revilla, J., Dubied, M., Montillot, C., Wentworth, E., Poloni, L., Varón-González, C., Jones, P., Xiong, Z., Fuentes-Guajardo, M., Palmal, S., Chacón-Duque, J. C., Hurtado, M., Villegas, V., Granja, V., Jaramillo, C., Arias, W., … Ruiz-Linares, A. (2021). A GWAS in Latin Americans identifies novel face shape loci, implicating VPS13B and a Denisovan introgressed region in facial variation. Science Advances, 7(6), eabc6160. 10.1126/sciadv.abc6160

9. Butali, A., Mossey, P. A., Adeyemo, W. L., Eshete, M. A., Gowans, L. J. J., Busch, T. D., Jain, D., Yu, W., Huan, L., Laurie, C. A., Laurie, C. C., Nelson, S., Li, M., Sanchez-Lara, P. A., Magee, W. P., Magee, K. S., Auslander, A., Brindopke, F., Kay, D. M., … Adeyemo, A. A. (2019). Genomic analyses in African populations identify novel risk loci for cleft palate. Human Molecular Genetics, 28(6), 1038–1051. 10.1093/hmg/ddy402

10. Carlson, J. C., & Leslie, E. J. (2021). The PAX1 locus at 20p11 is a potential genetic modifier for bilateral cleft lip. HGG Advances, 2(2), 100025. 10.1016/j.xhgg.2021.100025

11. Castanet, M., Park, S. M., Smith, A., Bost, M., Léger, J., Lyonnet, S., Pelet, A., Czernichow, P., Chatterjee, K., & Polak, M. (2002). A novel loss-of-function mutation in TTF-2 is associated with congenital hypothyroidism, thyroid agenesis, and cleft palate. Human Molecular Genetics, 11(17), 2051–2059. 10.1093/hmg/11.17.2051

12. Cha, S., Lim, J. E., Park, A. Y., Do, J. H., Lee, S. W., Shin, C., Cho, N. H., Kang, J. O., Nam, J. M., Kim, J. S., Woo, K. M., Lee, S. H., Kim, J. Y., & Oh, B. (2018). Identification of five novel genetic loci related to facial morphology by genome-wide association studies. BMC Genomics, 19(1), 481. 10.1186/s12864-018-4865-9

13. Cha, M. Y., Hong, Y. J., Choi, J. E., Kwon, T. S., Kim, I. J., & Hong, K. W. (2022). Classification of early age facial growth pattern and identification of the genetic basis in two Korean populations. Scientific Reports, 12(1), 13828. 10.1038/s41598-022-18127-6

14. Cho, H. W., Ban, H. J., Jin, H. S., Cha, S., & Eom, Y. B. (2023). Effect of genetic variants in UBE2O and TPK1 on facial morphology of Koreans. Forensic Science Research, 8(1), 62– 69. 10.1093/fsr/owad011

15. Curtis, S. W., Chang, D., Sun, M. R., Epstein, M. P., Murray, J. C., Feingold, E., Beaty, T. H., Weinberg, S. M., Marazita, M. L., Lipinski, R. J., Carlson, J. C., & Leslie, E. J. (2021). FAT4 identified as a potential modifier of orofacial cleft laterality. Genetic Epidemiology, 45(7), 721–735. 10.1002/gepi.22420

16. Curtis, S. W., Chang, D., Lee, M. K., Shaffer, J. R., Indencleef, K., Epstein, M. P., Cutler, D. J., Murray, J. C., Feingold, E., Beaty, T. H., Claes, P., Weinberg, S. M., Marazita, M. L., Carlson, J. C., & Leslie, E. J. (2021). The PAX1 locus at 20p11 is a potential genetic modifier for bilateral cleft lip. HGG Advances, 2(2), 100025. 10.1016/j.xhgg.2021.100025

17. Connelly, R., & Platt, L. (2014). Cohort profile: UK Millennium Cohort Study (MCS). International journal of epidemiology, 43(6), 1719–1725. 10.1093/ije/dyu001

18. da Silva Franco, J. F., Neves, R. L., Menezes, A. N., Nogueira, B. R., Gomes, C. P., & Pesquero, J. B. (2025). Hidden Aberrant Transcripts in TTC37 Cause Trichohepatoenteric Syndrome. Clinical genetics, 107(1), 113–114. 10.1111/cge.14630

19. Davies, A. J. V., Humphries, K., Lewis, S. J., Ho, K., Sandy, J. R., & Wren, Y. (2024). The Cleft Collective: protocol for a longitudinal prospective cohort study. BMJ open, 14(7), e084737. 10.1136/bmjopen-2024-084737

20. Davies, A. J. V., Wren, Y. E., Hamilton, M., Sandy, J. R., Stergiakouli, E., & Lewis, S. J. (2024). Predicting Syndromic Status Based on Longitudinal Data from Parental Reports of the Presence of Additional Structural and Functional Anomalies in Children Born with an Orofacial Cleft. Journal of clinical medicine, 13(22), 6924. 10.3390/jcm13226924

21. Demeer, B., Revencu, N., Helaers, R., Gbaguidi, C., Dakpe, S., Francois, G., Devauchelle, B., Bayet, B. and Vikkula, M. (2019) Likely pathogenic variants in one third of non-syndromic discontinuous cleft lip and palate patients. Genes (Basel), 10, 833.

22. Dixon, M. J., Marazita, M. L., Beaty, T. H., & Murray, J. C. (2011). Cleft lip and palate: understanding genetic and environmental influences. Nature reviews. Genetics, 12(3), 167– 178. 10.1038/nrg2933

23. Fairchild, C. L., & Gammill, L. S. (2013). Tetraspanin18 is a FoxD3-responsive antagonist of cranial neural crest epithelial-to-mesenchymal transition that maintains cadherin-6B protein. Journal of Cell Science, 126(6), 1464–1476. 10.1242/jcs.120915

24. Fitzsimons, E., Moulton, V., Hughes, D. A., Neaves, S., Ho, K., Hemani, G., Timpson, N., Calderwood, L., Gilbert, E., & Ring, S. (2022). Collection of genetic data at scale for a nationally representative population: The UK Millennium Cohort Study. Longitudinal and Life Course Studies, 13(1), 169–187. 10.1332/175795921X16334229321103

25. Fishilevich, S., Nudel, R., Rappaport, N., Hadar, R., Plaschkes, I., Iny Stein, T., Rosen, N., Kohn, A., Twik, M., Safran, M., Lancet, D., & Cohen, D. (2017). GeneHancer: Genome-wide integration of enhancers and target genes in GeneCards. Database, 2017, bax028. 10.1093/database/bax028

26. Gangopadhyay, N., Mendonca, D. A., & Woo, A. S. (2012). Pierre Robin sequence. Seminars in Plastic Surgery, 26(2), 76–82. 10.1055/s-0032-1320065

27. Grant, S. F., Wang, K., Zhang, H., Glaberson, W., Annaiah, K., Kim, C. E., Bradfield, J. P., Glessner, J. T., Thomas, K. A., Garris, M., Frackelton, E. C., Otieno, F. G., Chiavacci, R. M., Nah, H. D., Kirschner, R. E., & Hakonarson, H. (2009). A genome-wide association study identifies a locus for nonsyndromic cleft lip with or without cleft palate on 8q24. The Journal of Pediatrics, 155(6), 909–913. 10.1016/j.jpeds.2009.06.020

28. Gómez-Valdés, J., Villamil-Ramírez, H., Silva de Cerqueira, C. C., Hünemeier, T., Ramallo, V., Liu, F., Weinberg, S. M., Shaffer, J. R., Stergiakouli, E., Howe, L. J., Hysi, P. G., Spector, T. D., Gonzalez-José, R., Schüler-Faccini, L., Bortolini, M. C., Acuña-Alonzo, V., Canizales-Quinteros, S., Gallo, C., Poletti, G., … Ruiz-Linares, A. (2021). A GWAS in Latin Americans identifies novel face shape loci, implicating VPS13B and a Denisovan introgressed region in facial variation. Science Advances, 7(6), eabc6160. 10.1126/sciadv.abc6160

29. Gossen, S., Gerstner, S., & Borchers, A. (2024). The RhoGEF Trio is transported by microtubules and affects microtubule stability in migrating neural crest cells. Cells & development, 177, 203899. 10.1016/j.cdev.2023.203899

30. Hemani, G., Elsworth, B., Palmer, T., & Rasteiro, R. (2024). ieugwasr: Interface to the ’OpenGWAS’ Database API. R package version 1.0.1. https://github.com/MRCIEU/ieugwasr

31. Hoskens, H., Liu, D., Naqvi, S., Lee, M. K., Eller, R. J., Indencleef, K., White, J. D., Li, J., Larmuseau, M. H. D., Hens, G., Wysocka, J., Walsh, S., Richmond, S., Shriver, M. D., Shaffer, J. R., Peeters, H., Weinberg, S. M., & Claes, P. (2021). 3D facial phenotyping by biometric sibling matching used in contemporary genomic methodologies. PLoS genetics, 17(5), e1009528. 10.1371/journal.pgen.1009528

32. Huang, L., Jia, Z., Shi, Y., Du, Q., Shi, J., Wang, Z., Mou, Y., Wang, Q., Zhang, B., Wang, Q., et al. (2019). Genetic factors define CPO and CLO subtypes of nonsyndromic orofacial cleft. PLoS Genetics, 15(10), e1008357. 10.1371/journal.pgen.1008357

33. Huang, Q.Q., Wigdor, E.M., Malawsky, D.S. et al. Examining the role of common variants in rare neurodevelopmental conditions. Nature 636, 404–411 (2024). 10.1038/s41586-024-08217-yImuta

34. Y., Nishioka, N., Kiyonari, H., & Sasaki, H. (2009). Short limbs, cleft palate, and delayed formation of flat proliferative chondrocytes in mice with targeted disruption of a putative protein kinase gene, Pkdcc (AW548124). Developmental Dynamics, 238(1), 210– 222. 10.1002/dvdy.21822

35. Indencleef, K., Hoskens, H., Lee, M. K., White, J. D., Liu, C., Eller, R. J., Naqvi, S., Wehby, G. L., Moreno Uribe, L. M., Hecht, J. T., Long, R. E., Jr, Christensen, K., Deleyiannis, F. W., Walsh, S., Shriver, M. D., Richmond, S., Wysocka, J., Peeters, H., Shaffer, J. R., Marazita, M. L., … Claes, P. (2021). The Intersection of the Genetic Architectures of Orofacial Clefts and Normal Facial Variation. Frontiers in genetics, 12, 626403. 10.3389/fgene.2021.626403

36. Jia Z, Mukhopadhyay N, Yang Z, Butali A, Sun J, You Y, Yao M, Zhen Q, Ma J, He M, Pan Y, Alade A, Wang Y, Olujitan M, Qi M, Adeyemo WL, Buxó CJ, Gowans LJJ, Eshete M, Huang Y, Li C, Leslie EJ, Wang L, Bian Z, Carlson JC, Shi B, Weinberg SM, Murray JC, Sun L, Marazita ML, Freathy RM, Beaumont RN. Multi-ancestry Genome Wide Association Study Meta-analysis of Non-syndromic Orofacial Clefts. medRxiv [Preprint]. 2024 Dec 13:2024.12.06.24318522. doi: 10.1101/2024.12.06.24318522.

37. Kent, O. A., Saha, M., Coyaud, E., Burston, H. E., Law, N., Dadson, K., Chen, S., Laurent, E. M., St-Germain, J., Sun, R. X., Matsumoto, Y., Cowen, J., Montgomery-Song, A., Brown, K. R., Ishak, C., Rose, J., De Carvalho, D. D., He, H. H., Raught, B., Billia, F., … Rottapel, R. (2020). Haploinsufficiency of RREB1 causes a Noonan-like RASopathy via epigenetic reprogramming of RAS-MAPK pathway genes. Nature communications, 11(1), 4673. 10.1038/s41467-020-18483-9

38. Khan, M. I., Cs, P., & Srinath, N. (2022). Role of PAX7 gene rs766325 and rs4920520 polymorphisms in the etiology of non-syndromic cleft lip and palate: A genetic study. Global Medical Genetics, 9(3), 208–211. 10.1055/s-0042-1748531

39. Khan, M. I., Cs, P., Mustak, M. S., & Nizamuddin, S. (2023). Maternal transmission of the PAX7 single nucleotide polymorphisms among Indian cleft trios. Global Medical Genetics, 10(1), 6–11. 10.1055/s-0042-1760383

40. Knapp, M. (1999). A note on power approximations for the transmission/disequilibrium test. American Journal of Human Genetics, 64, 1177–1185.

41. Lee, J. J., McGue, M., Iacono, W. G., & Chow, C. C. (2018). The accuracy of LD Score regression as an estimator of confounding and genetic correlations in genome-wide association studies. Genetic epidemiology, 42(8), 783–795. 10.1002/gepi.22161

42. Lee, M. K., Shaffer, J. R., Leslie, E. J., Orlova, E., Carlson, J. C., Feingold, E., Marazita, M. L., & Weinberg, S. M. (2017). Genome-wide association study of facial morphology reveals novel associations with FREM1 and PARK2. PLoS One, 12(4), e0176566. 10.1371/journal.pone.0176566

43. Leslie, E. J., & Marazita, M. L. (2013). Genetics of cleft lip and cleft palate. American journal of medical genetics. Part C, Seminars in medical genetics, *163C*(4), 246–258. 10.1002/ajmg.c.31381

44. Leslie, E. J., Liu, H., Carlson, J. C., Shaffer, J. R., Feingold, E., Wehby, G., Laurie, C. A., Jain, D., Laurie, C. C., Doheny, K. F., McHenry, T., Resick, J., Sanchez, C., Jacobs, J., Emanuele, B., Vieira, A. R., Neiswanger, K., Standley, J., Czeizel, A. E., … Marazita, M. L. (2016). A genome-wide association study of nonsyndromic cleft palate identifies an etiologic missense variant in GRHL3. The American Journal of Human Genetics, 98(4), 744–754. 10.1016/j.ajhg.2016.02.014

45. Leslie, E. J., Carlson, J. C., Shaffer, J. R., Butali, A., Buxó, C. J., Castilla, E. E., Christensen, K., Deleyiannis, F. W., Field, L. L., Hecht, J. T., Moreno, L., Orioli, I. M., Padilla, C., Vieira, A. R., Wehby, G. L., Feingold, E., Weinberg, S. M., Murray, J. C., Beaty, T. H., & Marazita, M. L. (2017). Genome-wide meta-analyses of nonsyndromic orofacial clefts identify novel associations between FOXE1 and all orofacial clefts, and TP63 and cleft lip with or without cleft palate. Human Genetics, 136(3), 275–286. 10.1007/s00439-016-1754-7

46. Li, Q., Chen, J., Faux, P., Delgado, M. E., Bonfante, B., Fuentes-Guajardo, M., Mendoza-Revilla, J., Chacón-Duque, J. C., Hurtado, M., Villegas, V., Granja, V., Jaramillo, C., Arias, W., Barquera, R., Everardo-Martínez, P., Sánchez-Quinto, M., Gómez-Valdés, J., Villamil-Ramírez, H., Silva de Cerqueira, C. C., Hünemeier, T., … Ruiz-Linares, A. (2023). Automatic landmarking identifies new loci associated with face morphology and implicates Neanderthal introgression in human nasal shape. Communications biology, 6(1), 481. 10.1038/s42003-023-04838-7

47. Lim, K. C., Lakshmanan, G., Crawford, S. E., Gu, Y., Grosveld, F., & Engel, J. D. (2000). Gata3 loss leads to embryonic lethality due to noradrenaline deficiency of the sympathetic nervous system. Nature Genetics, 25(2), 209–212. 10.1038/76080

48. Ludwig, K. U., Mangold, E., Herms, S., Nowak, S., Reutter, H., Paul, A., Becker, J., Herberz, R., AIChawa, T., Nasser, E., et al. (2012). Genome-wide meta-analyses of nonsyndromic cleft lip with or without cleft palate identify six new risk loci. Nature Genetics, 44, 968–971. 10.1038/ng.2360

49. Ludwig, K. U., Böhmer, A. C., Rubini, M., Mossey, P. A., Herms, S., Nowak, S., Reutter, H., Alblas, M. A., Lippke, B., Barth, S., Paredes-Zenteno, M., Muñoz-Jimenez, S. G., Ortiz-Lopez, R., Kreusch, T., Hemprich, A., Martini, M., Braumann, B., Jäger, A., Pötzsch, B., Molloy, A., … Mangold, E. (2014). Strong association of variants around FOXE1 and orofacial clefting. Journal of dental research, 93(4), 376–381. 10.1177/0022034514523987

50. Ludwig, K. U., Böhmer, A. C., Bowes, J., Nikolic, M., Ishorst, N., Wyatt, N., Hammond, N. L., Gölz, L., Thieme, F., Barth, S., Schuenke, H., Klamt, J., Spielmann, M., Aldhorae, K., Rojas-Martinez, A., Nöthen, M. M., Rada-Iglesias, A., Dixon, M. J., Knapp, M., & Mangold, E. (2017). Imputation of orofacial clefting data identifies novel risk loci and sheds light on the genetic background of cleft lip ± cleft palate and cleft palate only. Human Molecular Genetics, 26(4), 829–842. 10.1093/hmg/ddx012

51. Ludwig, K. U., Böhmer, A. C., Bowes, J., Nikolic, M., Ishorst, N., Wyatt, N., Hammond, N. L., Gölz, L., Thieme, F., Barth, S., Schuenke, H., Klamt, J., Spielmann, M., Aldhorae, K., Rojas-Martinez, A., Nöthen, M. M., Rada-Iglesias, A., Dixon, M. J., Knapp, M., & Mangold, E. (2017). Imputation of orofacial clefting data identifies novel risk loci and sheds light on the genetic background of cleft lip ± cleft palate and cleft palate only. Human Molecular Genetics, 26(4), 829–842. 10.1093/hmg/ddx012

52. Liu, H., Leslie, E. J., Carlson, J. C., Beaty, T. H., Marazita, M. L., Lidral, A. C., & Cornell, R. A. (2017). Identification of common non-coding variants at 1p22 that are functional for non-syndromic orofacial clefting. Nature communications, 8, 14759. 10.1038/ncomms14759

53. Mangold, E., Ludwig, K. U., Birnbaum, S., Baluardo, C., Ferrian, M., Herms, S., Reutter, H., de Assis, N. A., Chawa, T. A., Mattheisen, M., Steffens, M., Barth, S., Kluck, N., Paul, A., Becker, J., Lauster, C., Schmidt, G., Braumann, B., Scheer, M., Reich, R. H., … Nöthen, M. M. (2010). Genome-wide association study identifies two susceptibility loci for nonsyndromic cleft lip with or without cleft palate. Nature genetics, 42(1), 24–26. 10.1038/ng.506

54. Meng, T., Shi, J. Y., Wu, M., Wang, Y., Li, L., Liu, Y., Zheng, Q., Huang, L., & Shi, B. (2012). Overexpression of mouse TTF-2 gene causes cleft palate. Journal of cellular and molecular medicine, 16(10), 2362–2368. 10.1111/j.1582-4934.2012.01546.x

55. Mossey, P. A., Little, J., Munger, R. G., Dixon, M. J., & Shaw, W. C. (2009). Cleft lip and palate. *Lancet (London*, England), 374(9703), 1773–1785. 10.1016/S0140-6736(09)60695-4

56. Mukhopadhyay, N., Feingold, E., Moreno-Uribe, L., Wehby, G., Valencia-Ramirez, L. C., Muñeton, C. P. R., Padilla, C., Deleyiannis, F., Christensen, K., Poletta, F. A., Orioli, I. M., Hecht, J. T., Buxó, C. J., Butali, A., Adeyemo, W. L., Vieira, A. R., Shaffer, J. R., Murray, J. C., Weinberg, S. M., Leslie, E. J., … Marazita, M. L. (2021). Genome-Wide Association Study of Non-syndromic Orofacial Clefts in a Multiethnic Sample of Families and Controls Identifies Novel Regions. Frontiers in cell and developmental biology, 9, 621482. 10.3389/fcell.2021.621482

57. Naqvi, S., Kim, S., Hoskens, H., Matthews, H. S., Spritz, R. A., Klein, O. D., Hallgrímsson, B., Swigut, T., Claes, P., Pritchard, J. K., & Wysocka, J. (2023). Precise modulation of transcription factor levels identifies features underlying dosage sensitivity. Nature genetics, 55(5), 841–851. 10.1038/s41588-023-01366-2

58. Paganini, C., Gramegna Tota, C., Superti-Furga, A., & Rossi, A. (2020). Skeletal Dysplasias Caused by Sulfation Defects. International journal of molecular sciences, 21(8), 2710. 10.3390/ijms21082710

59. Pakvasa, M., Haravu, P., Boachie-Mensah, M., Jones, A., Coalson, E., Liao, J., Zeng, Z., Wu, D., Qin, K., Wu, X., Luo, H., Zhang, J., Zhang, M., He, F., Mao, Y., Zhang, Y., Niu, C., Wu, M., Zhao, X., Wang, H., … Reid, R. R. (2020). Notch signaling: Its essential roles in bone and craniofacial development. Genes & diseases, 8(1), 8–24. 10.1016/j.gendis.2020.04.006

60. Panamonta, V., Pradubwong, S., Panamonta, M., & Chowchuen, B. (2015). Global Birth Prevalence of Orofacial Clefts: A Systematic Review. Journal of the Medical Association of Thailand = Chotmaihet thangphaet, 98 *Suppl 7*, S11–S21.

61. Pickrell, J. K., Berisa, T., Liu, J. Z., Ségurel, L., Tung, J. Y., & Hinds, D. A. (2016). Detection and interpretation of shared genetic influences on 42 human traits. Nature Genetics, 48(7), 709–717. 10.1038/ng.3570

62. Purcell, S., Neale, B., Todd-Brown, K., Thomas, L., Ferreira, M., Bender, D., Maller, J., Sklar, P., de Bakker, P., Daly, M. J., et al. (2007). PLINK: A tool set for whole-genome and population-based linkage analyses. American Journal of Human Genetics, 81(3), 559–575. 10.1086/519795

63. Pruim, R. J., Welch, R. P., Sanna, S., Teslovich, T. M., Chines, P. S., Gliedt, T. P., Boehnke, M., Abecasis, G. R., & Willer, C. J. (2010). LocusZoom: regional visualization of genome-wide association scan results. *Bioinformatics (Oxford*, England), 26(18), 2336–2337. 10.1093/bioinformatics/btq419

64. Rahimov F, Nieminen P, Kumari P, Juuri E, Nikopensius T, Paraiso K, German J, Karvanen A, Kals M, Elnahas AG, Karjalainen J, Kurki M, Palotie A; FinnGen; Estonian Biobank Research Team; Heliövaara A, Esko T, Jukarainen S, Palta P, Ganna A, Patni AP, Mar D, Bomsztyk K, Mathieu J, Ruohola-Baker H, Visel A, Fakhouri WD, Schutte BC, Cornell RA, Rice DP. High incidence and geographic distribution of cleft palate in Finland are associated with the IRF6 gene. Nat Commun. 2024 Nov 6;15(1):9568. doi: 10.1038/s41467-024-53634-2.

65. Ray, D., Venkataraghavan, S., Zhang, W., et al. (2021). Pleiotropy method reveals genetic overlap between orofacial clefts at multiple novel loci from GWAS of multi-ethnic trios. PLOS Genetics, 17(7), e1009584. 10.1371/journal.pgen.1009584

66. Sanford, L. P., Ormsby, I., Gittenberger-de Groot, A. C., Sariola, H., Friedman, R., Boivin, G. P., Cardell, E. L., & Doetschman, T. (1997). TGFbeta2 knockout mice have multiple developmental defects that are non-overlapping with other TGFbeta knockout phenotypes. *Development (Cambridge*, England), 124(13), 2659–2670.

67. Sardiello, M., Annunziata, I., Roma, G., Ballabio, A. Sulfatases and sulfatase modifying factors: an exclusive and promiscuous relationship. Hum. Molec. Genet. 14: 3203–3217, 2005.

68. Selvi, R., & Mukunda, A. P. (2013). Role of SOX9 in the etiology of Pierre-Robin syndrome. Iranian Journal of Basic Medical Sciences, 16(5), 700–704.

69. Shaffer, J. R., Orlova, E., Lee, M. K., Leslie, E. J., Raffensperger, Z. D., Heike, C. L., … Weinberg, S. M. (2016). Genome-wide association study reveals multiple loci influencing normal human facial morphology. PLOS Genetics, 12(8), e1006149. 10.1371/journal.pgen.1006149

70. Sun, Y., Huang, Y., Yin, A., Pan, Y., Wang, Y., Wang, C., … Yang, Y. (2015). Genome-wide association study identifies a new susceptibility locus for cleft lip with or without a cleft palate. Nature Communications, 6, 6414. 10.1038/ncomms7414

71. Swartz, M. E., Lovely, C. B., & Eberhart, J. K. (2021). Variation in phenotypes from a Bmp- Gata3 genetic pathway is modulated by Shh signaling. PLoS Genetics, 17(5), e1009579. 10.1371/journal.pgen.1009579

72. Taliun, D., Harris, D. N., Kessler, M. D., et al. (2021). Sequencing of 53,831 diverse genomes from the NHLBI TOPMed Program. Nature, 590, 290–299. 10.1038/s41586-021-03205-y

73. Turner, S. (2018). qqman: An R package for visualizing GWAS results using Q-Q and manhattan plots. Journal of Open Source Software, 3(25), 731. 10.21105/joss.00731

74. Van Esch, H., Groenen, P., Nesbit, M. A., Schuffenhauer, S., Lichtner, P., Vanderlinden, G., Harding, B., Beetz, R., Bilous, R. W., Holdaway, I., Shaw, N. J., Fryns, J. P., Van de Ven, W., Thakker, R. V., & Devriendt, K. (2000). GATA3 haplo-insufficiency causes human HDR syndrome. Nature, 406(6794), 419–422. 10.1038/35019088

75. Varadarajan, S., Balaji, T. M., Raj, A. T., Gupta, A. A., Patil, S., Alhazmi, T. H., Alaqi, H. A. A., Al Omar, N. E. M., Almutaher, S. A. B. A., Jafer, A. A., et al. (2021). Genetic mutations associated with Pierre Robin syndrome/sequence: A systematic review. Molecular Syndromology, 12(2), 69–86. 10.1159/000513217

76. Verheyen, S., Blatterer, J., Speicher, M. R., Bhavani, G. S., Boons, G.-J., Ilse, M.-B., Andrae, D., Spross, J., Vaz, F. M., Kircher, S. G., Posch-Pertl, L., Baumgartner, D., Lubke, T., Shah, H., Al Kaissi, A., Girisha, K., Plecko, B. Novel subtype of mucopolysaccharidosis caused by arylsulfatase K (ARSK) deficiency. J. Med. Genet. 59: 957–964, 2022.

77. Vieira, A. R., Avila, J. R., Daack-Hirsch, S., Dragan, E., Félix, T. M., Rahimov, F., Harrington, J., Schultz, R. R., Watanabe, Y., Johnson, M., Fang, J., O’Brien, S. E., Orioli, I. M., Castilla, E. E., Fitzpatrick, D. R., Jiang, R., Marazita, M. L., & Murray, J. C. (2005). Medical sequencing of candidate genes for nonsyndromic cleft lip and palate. PLoS genetics, 1(6), e64. 10.1371/journal.pgen.0010064

78. Wang, P., Wu, Y., Li, Y., Zheng, J., & Tang, J. (2013). A novel RING finger E3 ligase RNF186 regulates ER stress-mediated apoptosis through interaction with BNip1. Cellular Signalling, 25(11), 2320–2333. 10.1016/j.cellsig.2013.07.016

79. Watanabe, K., Taskesen, E., van Bochoven, A., et al. (2017). Functional mapping and annotation of genetic associations with FUMA. Nature Communications, 8, 1826. 10.1038/s41467-017-01261-5

80. Welzenbach, J., Hammond, N. L., Nikolić, M., Thieme, F., Ishorst, N., Leslie, E. J., Weinberg, S. M., Beaty, T. H., Marazita, M. L., Mangold, E., Knapp, M., Cotney, J., Rada-Iglesias, A., Dixon, M. J., & Ludwig, K. U. (2021). Integrative approaches generate insights into the architecture of non-syndromic cleft lip with or without cleft palate. HGG advances, 2(3), 100038. 10.1016/j.xhgg.2021.100038

81. White, J. D., Indencleef, K., Naqvi, S., Eller, R. J., Hoskens, H., Roosenboom, J., Lee, M. K., Li, J., Mohammed, J., Richmond, S., Quillen, E. E., Norton, H. L., Feingold, E., Swigut, T., Marazita, M. L., Peeters, H., Hens, G., Shaffer, J. R., Wysocka, J., Walsh, S., … Claes, P. (2021). Insights into the genetic architecture of the human face. Nature genetics, 53(1), 45–53. 10.1038/s41588-020-00741-7

82. Wilson, K., Newbury, D. F., & Kini, U. (2023). Analysis of exome data in a UK cohort of 603 patients with syndromic orofacial clefting identifies causal molecular pathways. Human molecular genetics, 32(11), 1932–1942. 10.1093/hmg/ddad023

83. Xiong, Z., Dankova, G., Howe, L. J., Lee, M. K., Hysi, P. G., de Jong, M. A., Zhu, G., Adhikari, K., Li, D., Li, Y., Pan, B., Feingold, E., Marazita, M. L., Shaffer, J. R., McAloney, K., Xu, S. H., Jin, L., Wang, S., de Vrij, F. M., Lendemeijer, B., … International Visible Trait Genetics (VisiGen) Consortium (2019). Novel genetic loci affecting facial shape variation in humans. eLife, 8, e49898. 10.7554/eLife.49898

84. Yang, Y., Suzuki, A., Iwata, J., & Jun, G. (2020). Secondary Genome-Wide Association Study Using Novel Analytical Strategies Disentangle Genetic Components of Cleft Lip and/or Cleft Palate in 1q32.2. Genes, 11(11), 1280. 10.3390/genes11111280

85. Yu, Y., Zuo, X., He, M., Gao, J., Fu, Y., Qin, C., Meng, L., Wang, W., Song, Y., Cheng, Y., Zhou, F., Chen, G., Zheng, X., Wang, X., Liang, B., Zhu, Z., Fu, X., Sheng, Y., Hao, J., Liu, Z., … Bian, Z. (2017). Genome-wide analyses of non-syndromic cleft lip with palate identify 14 novel loci and genetic heterogeneity. Nature communications, 8, 14364. 10.1038/ncomms14364

86. Zhao, Y., Guo, Y. J., Tomac, A. C., Taylor, N. R., Grinberg, A., Lee, E. J., Huang, S., & Westphal, H. (1999). Isolated cleft palate in mice with a targeted mutation of the LIM homeobox gene lhx8. Proceedings of the National Academy of Sciences of the United States of America, 96(26), 15002–15006. 10.1073/pnas.96.26.15002

87. Zhu, B., Mandal, S. S., Pham, A. D., Zheng, Y., Erdjument-Bromage, H., Batra, S. K., Tempst, P., & Reinberg, D. (2005). The human PAF complex coordinates transcription with events downstream of RNA synthesis. Genes & Development, 19(14), 1668–1673. 10.1101/gad.1292105

