## Supplementary figures for "Genetic Heterogeneity and Homogeneity Among Orofacial Cleft Subtypes: Genome-Wide Association Studies in the Cleft Collective"

Supplementary Figure 1 – scatter plot showing principal component 1 versus principal component 2 for ancestry in cases versus controls

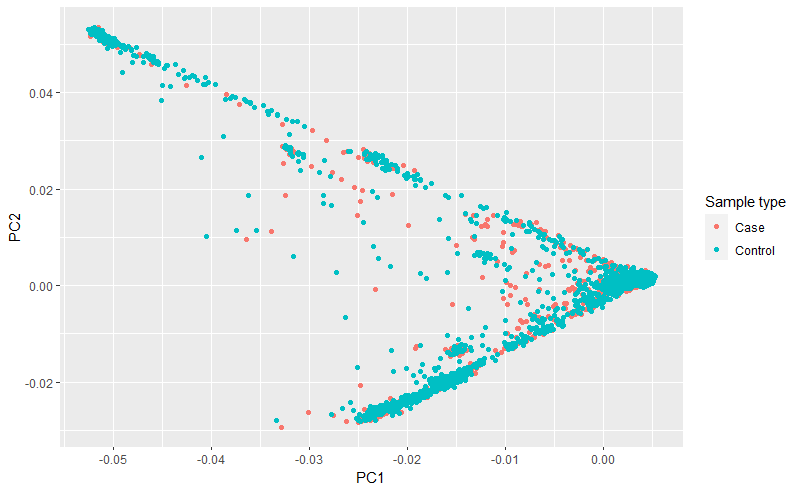

Supplementary Figure 2 – Manhattan plot – All cases

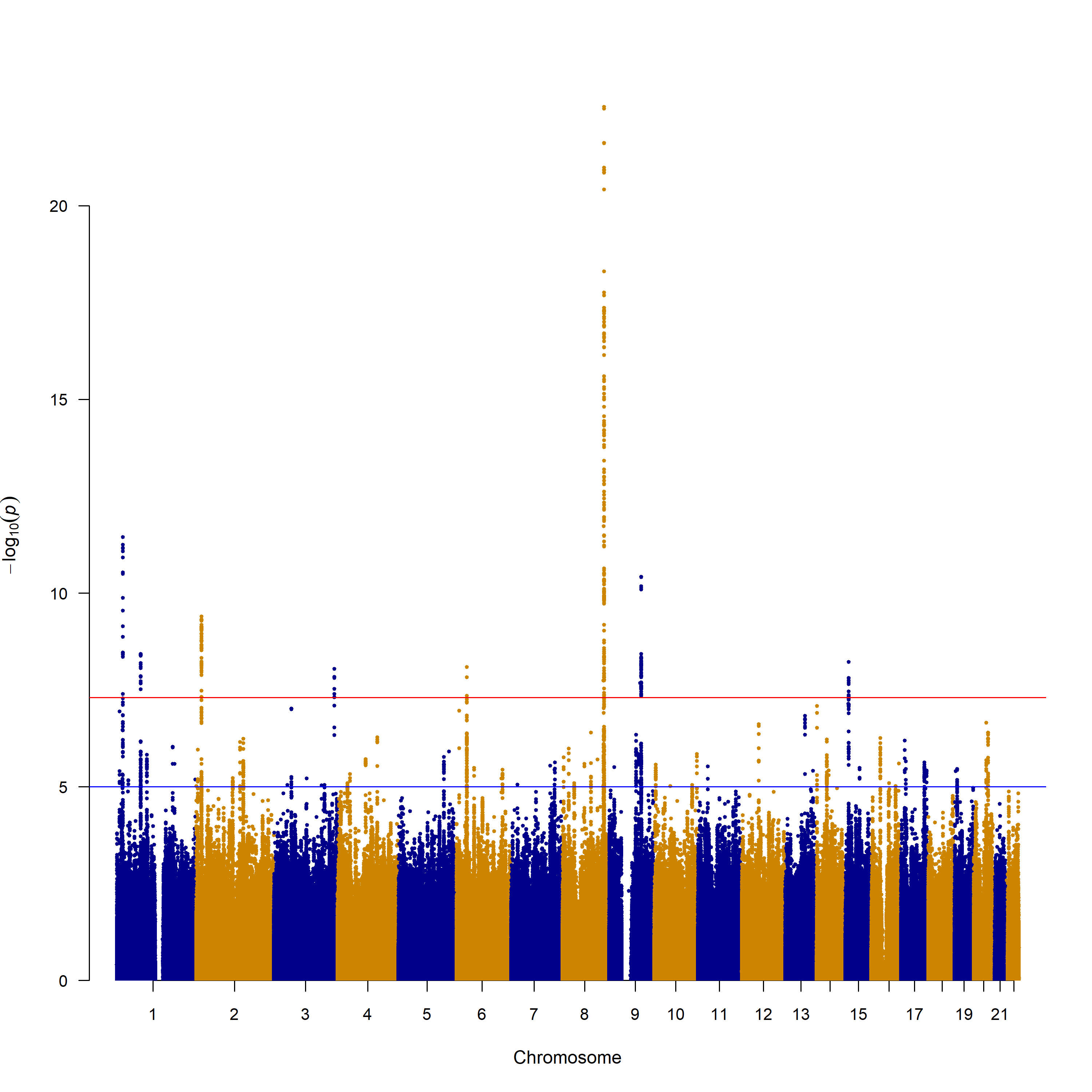

Supplementary Figure 3 – Quantile-Quantile plot – All cases

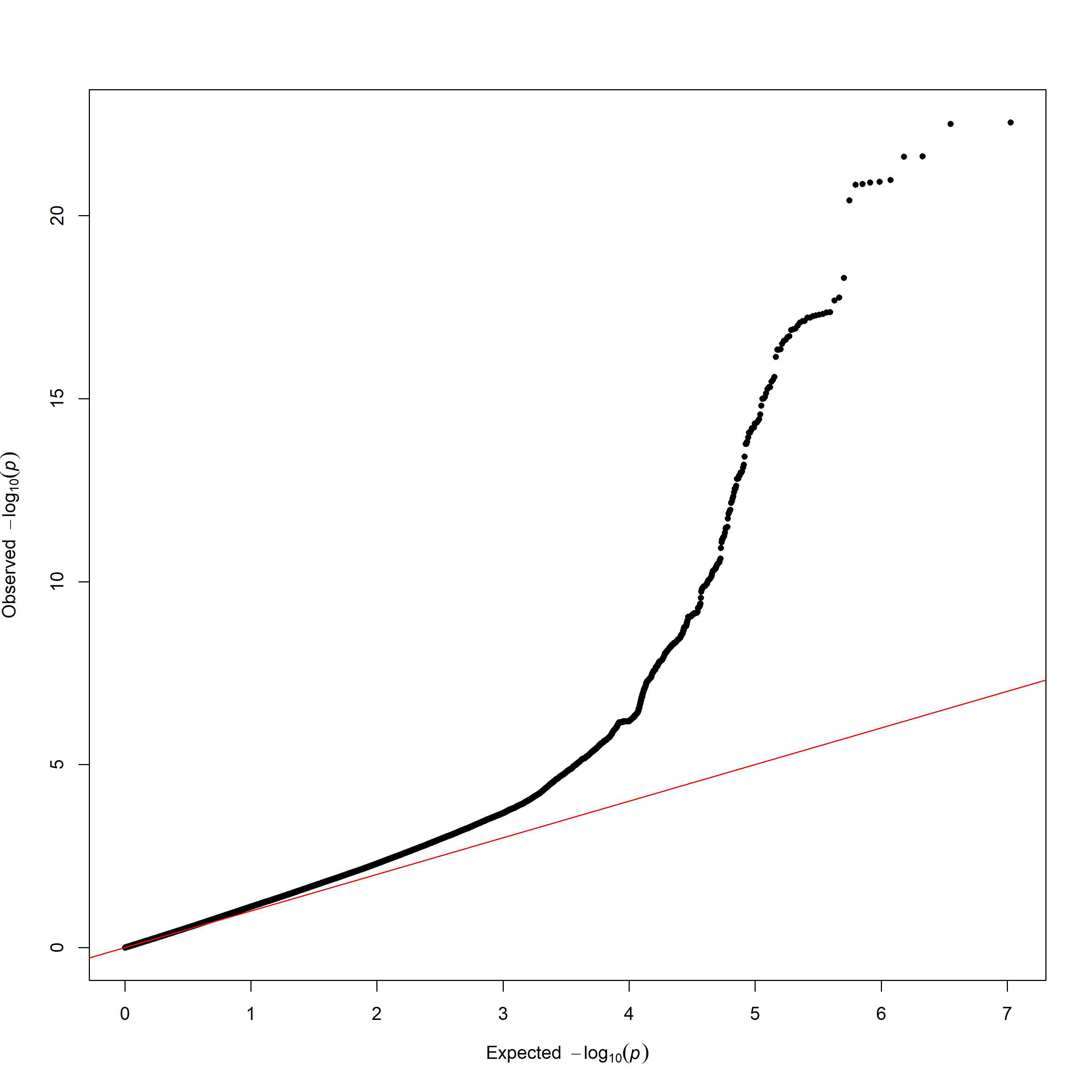

Supplementary Figure 4 – Manhattan plot – Cleft lip with or without cleft palate

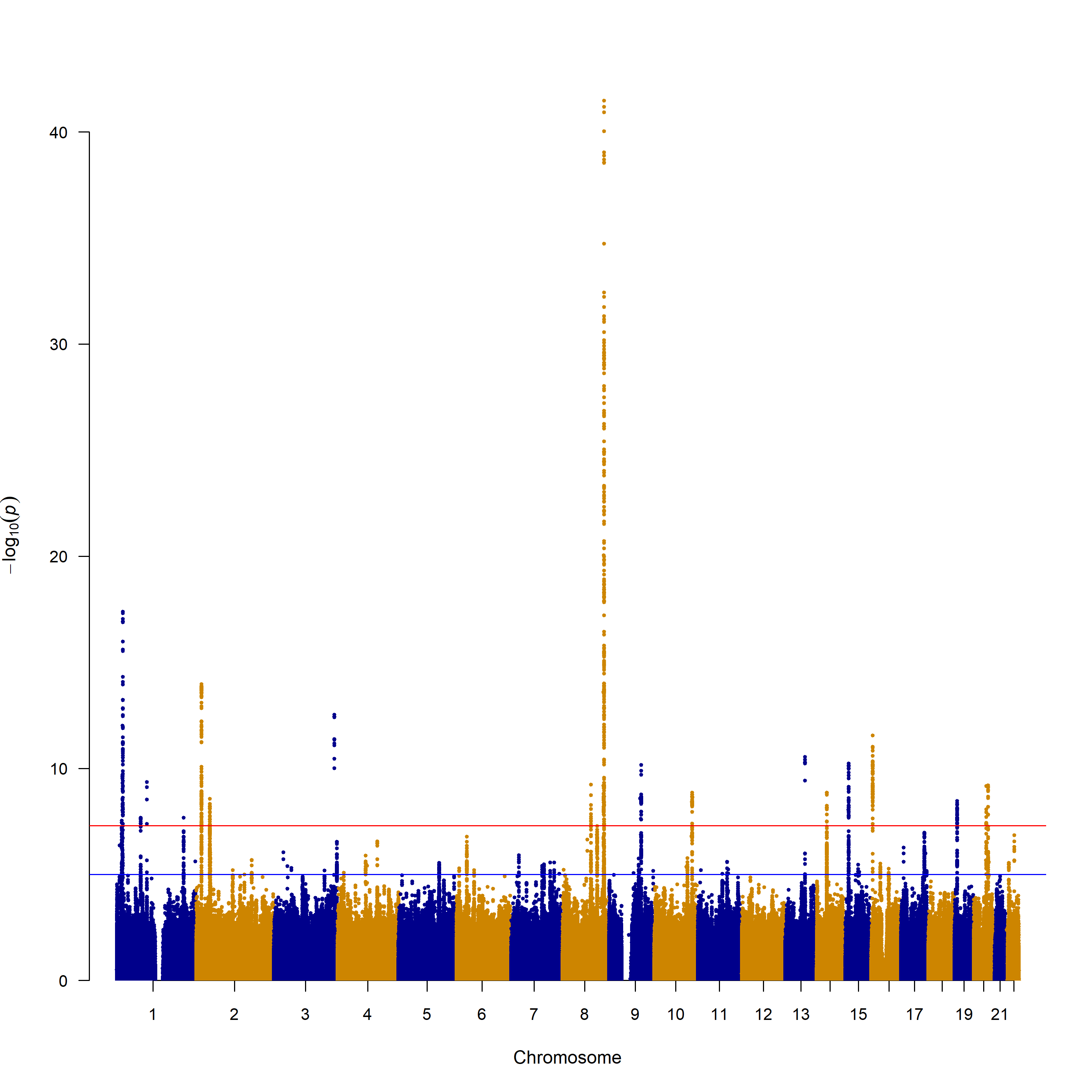

Supplementary Figure 5– Quantile-Quantile plot – Cleft lip with or without palate

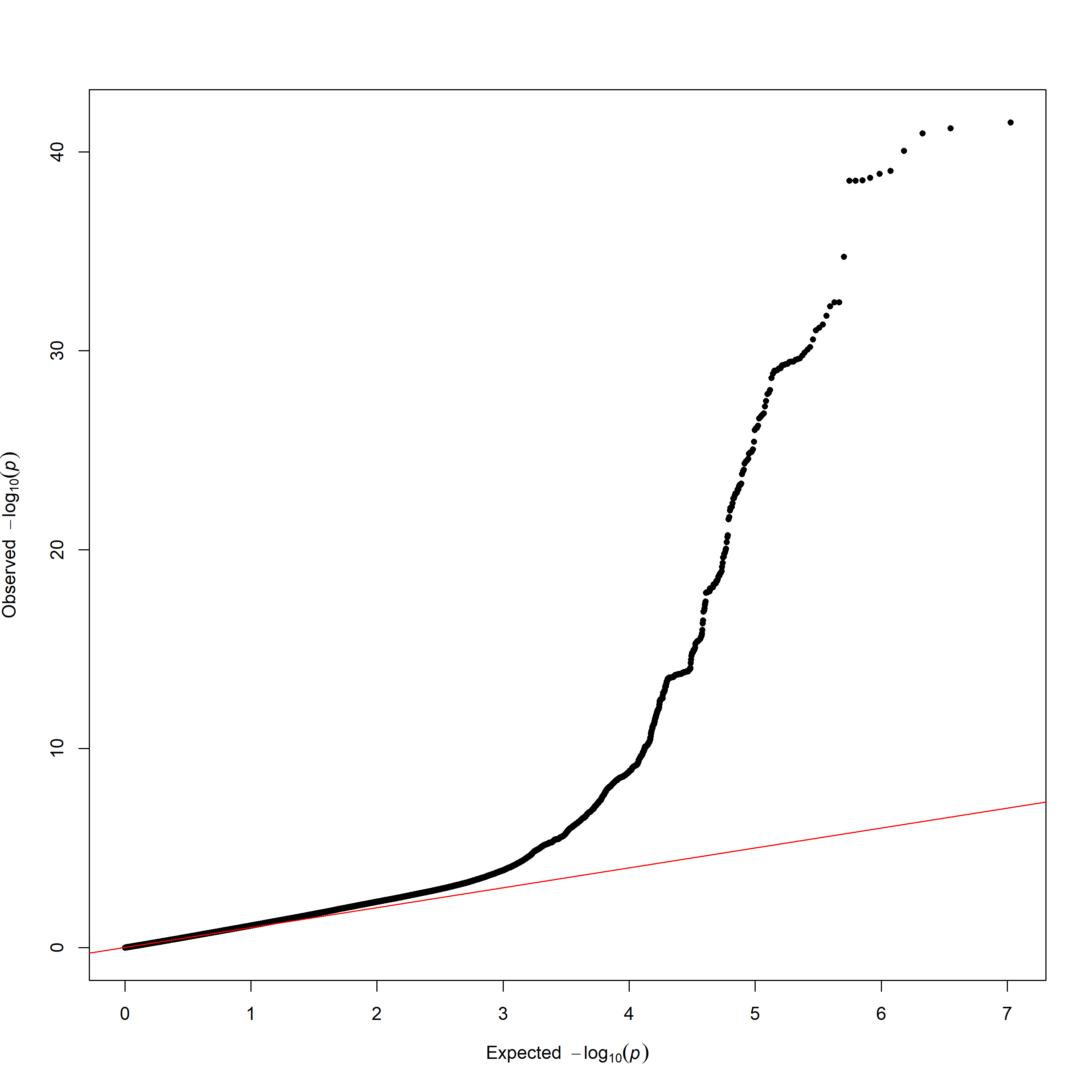

Supplementary Figure 6 – Manhattan plot – Non syndromic cleft lip with or without palate

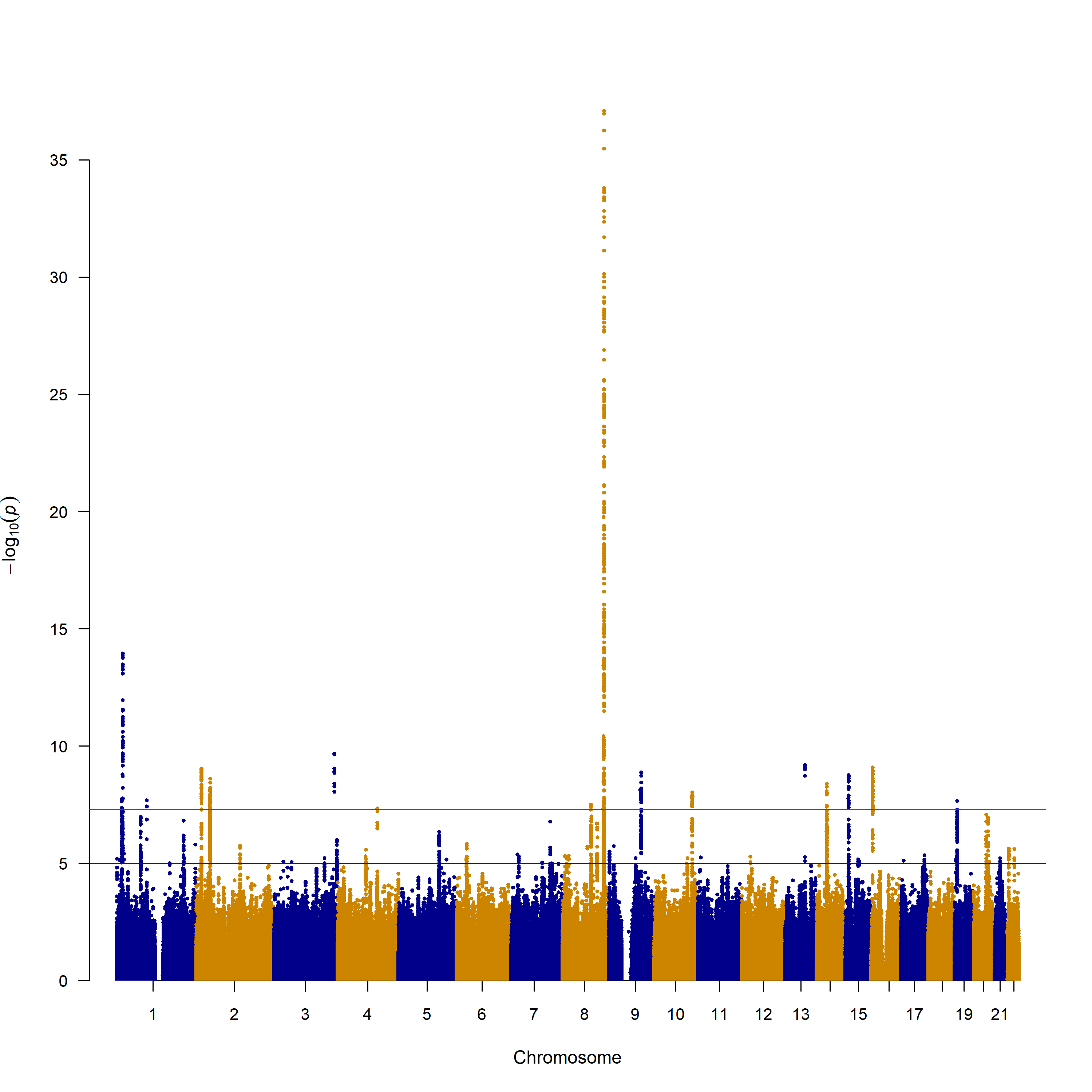

Supplementary Figure 7– Quantile-Quantile plot – Non syndromic cleft lip with or without palate

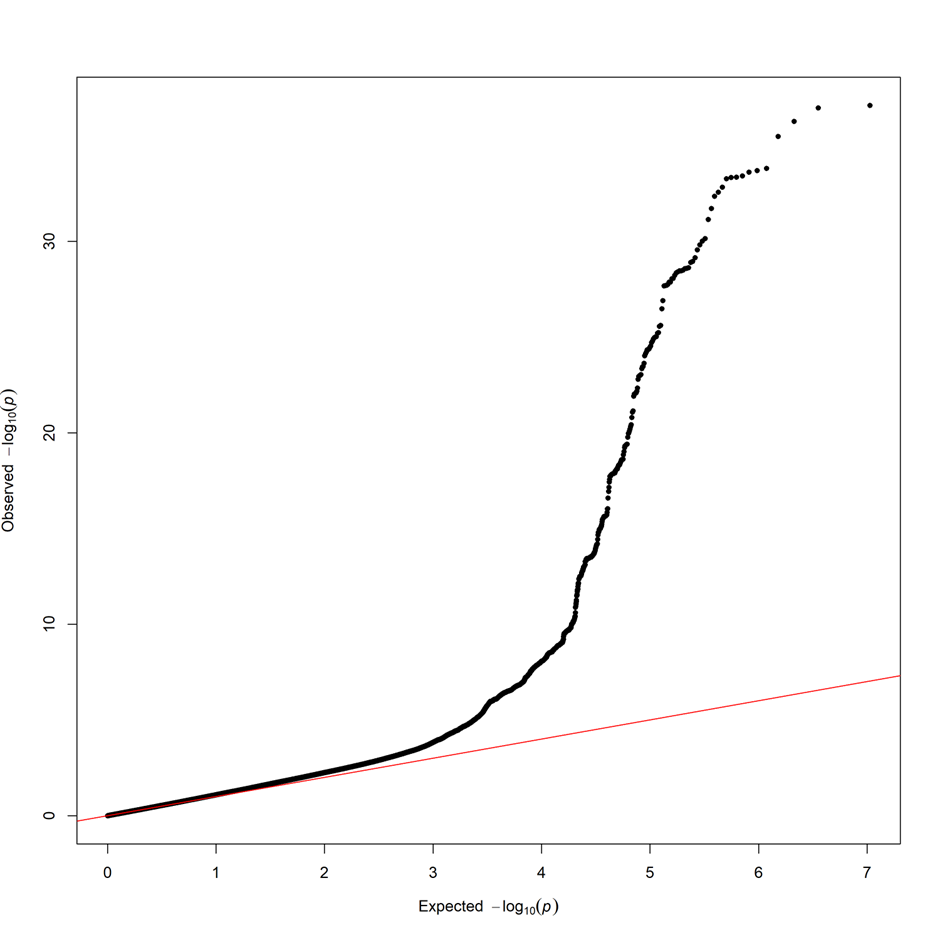

Supplementary Figure 8– Manhattan plot –Cleft lip with palate

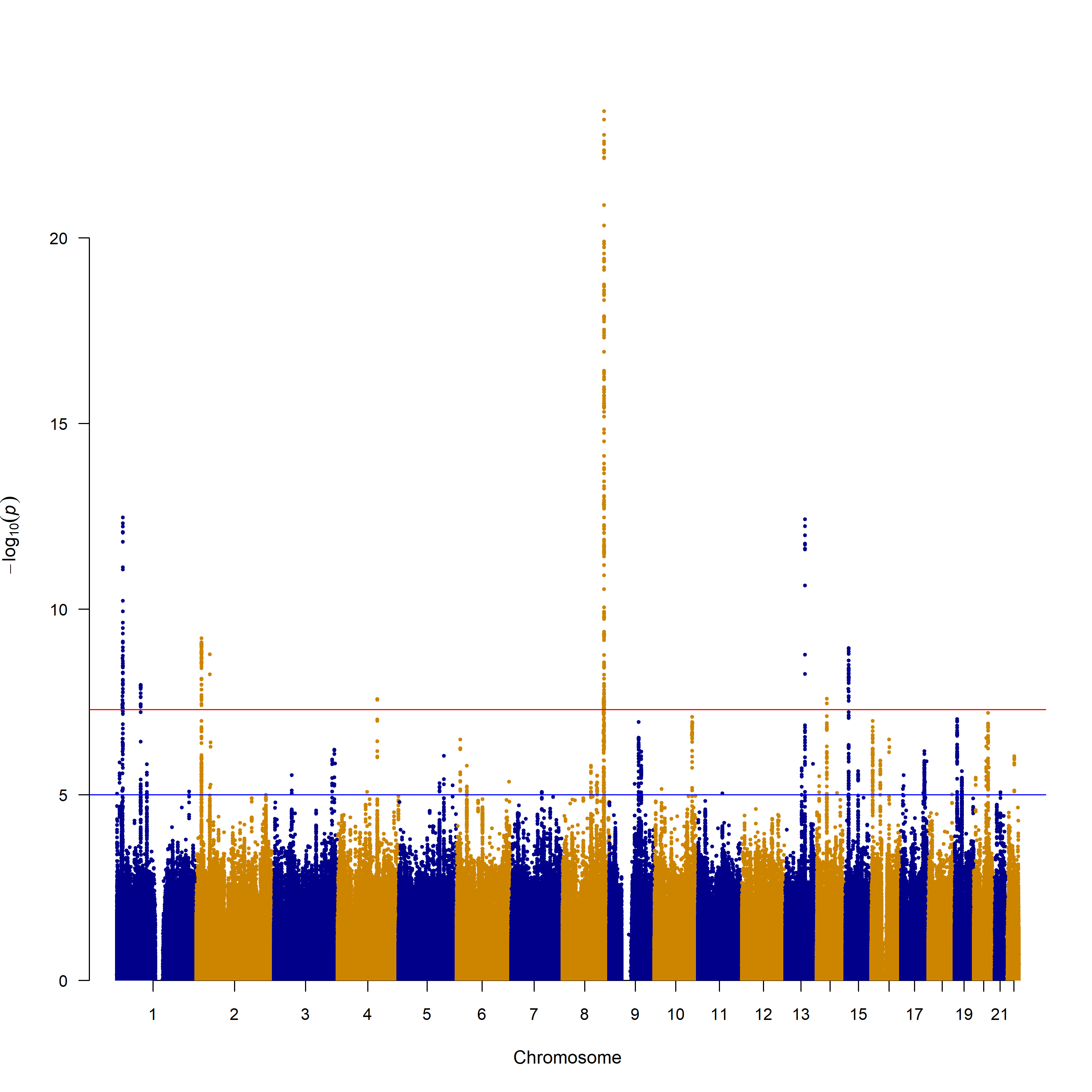

Supplementary Figure 9– Quantile-Quantile plot – Cleft lip with palate
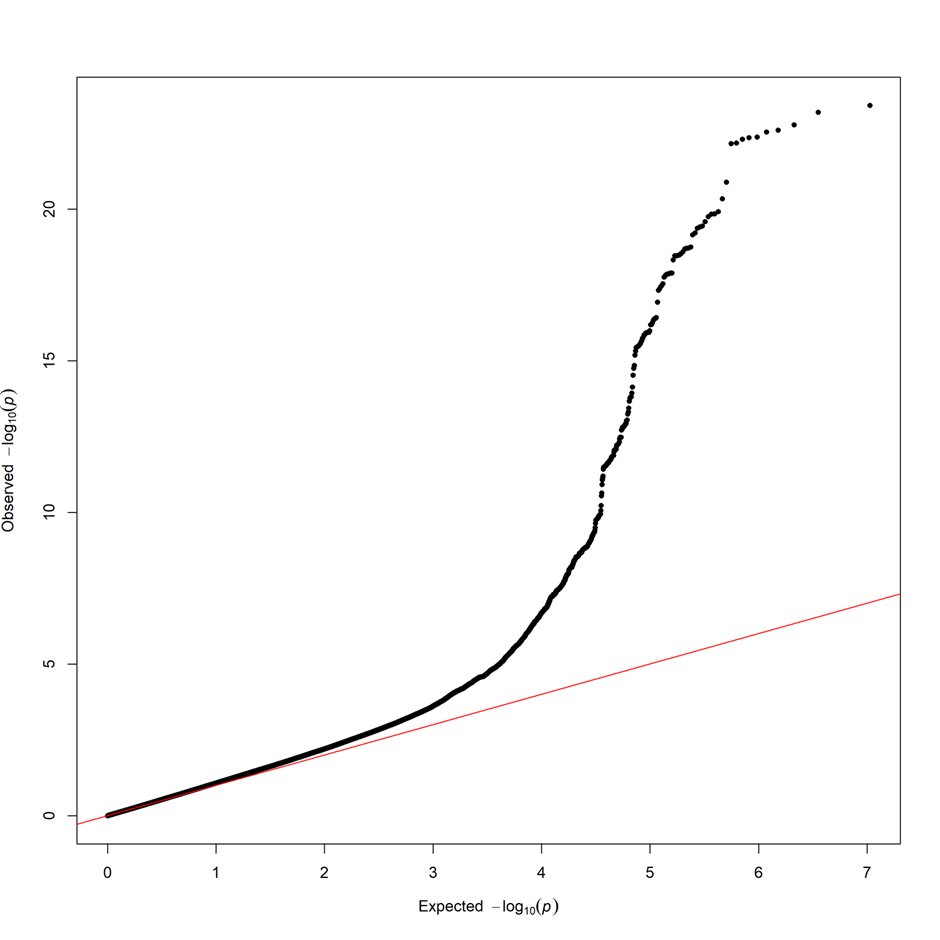

Supplementary Figure 10 – Manhattan plot –Cleft lip only

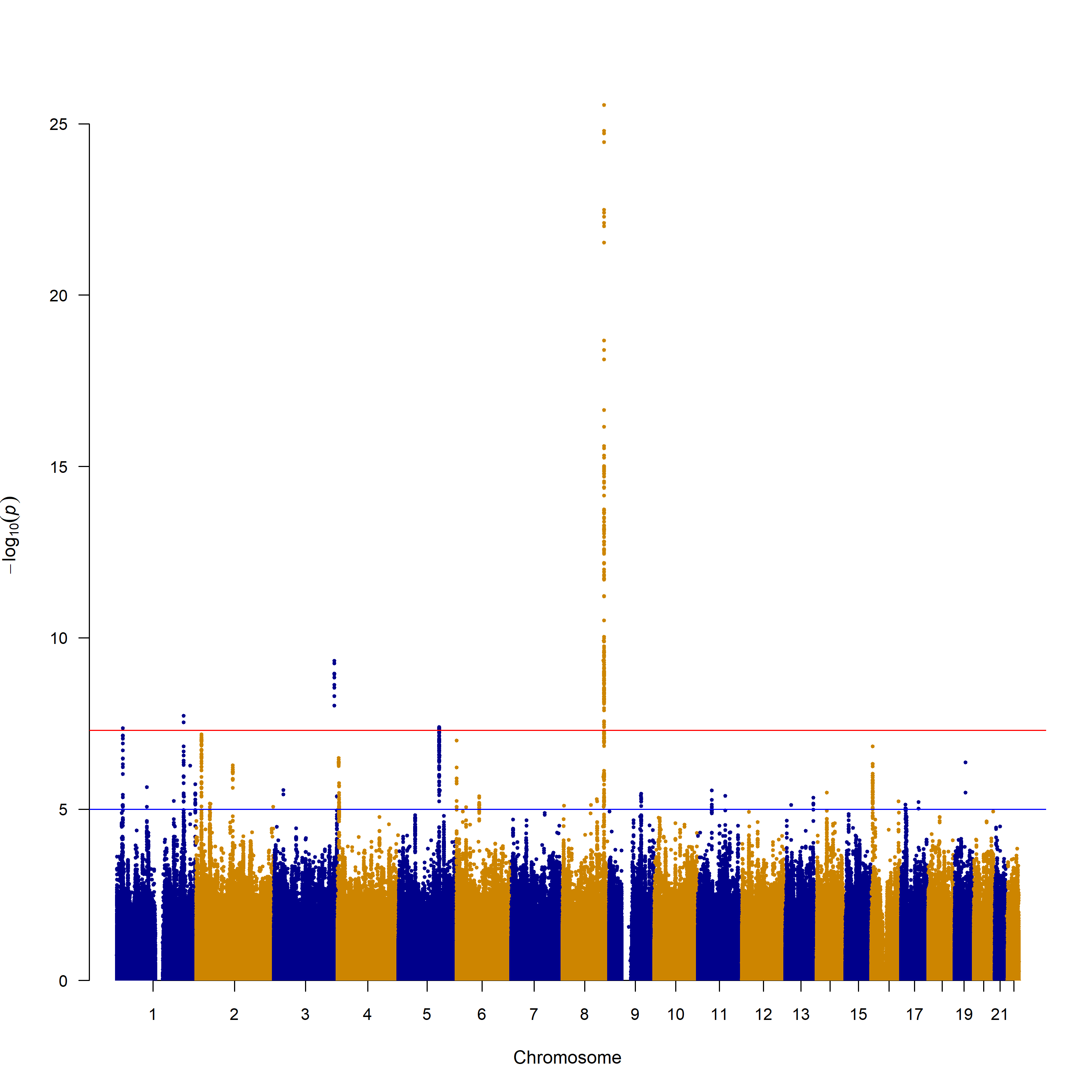

Supplementary Figure 11– Quantile-Quantile plot – Cleft lip only

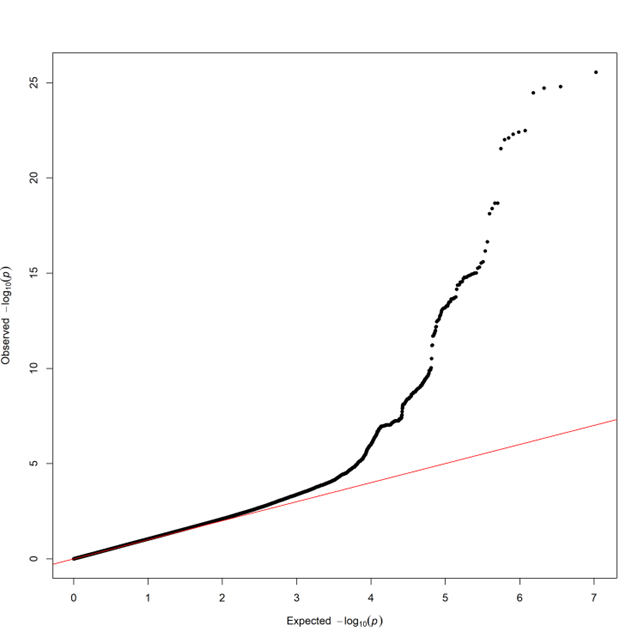

Supplementary Figure 12– Manhattan plot –Cleft palate only

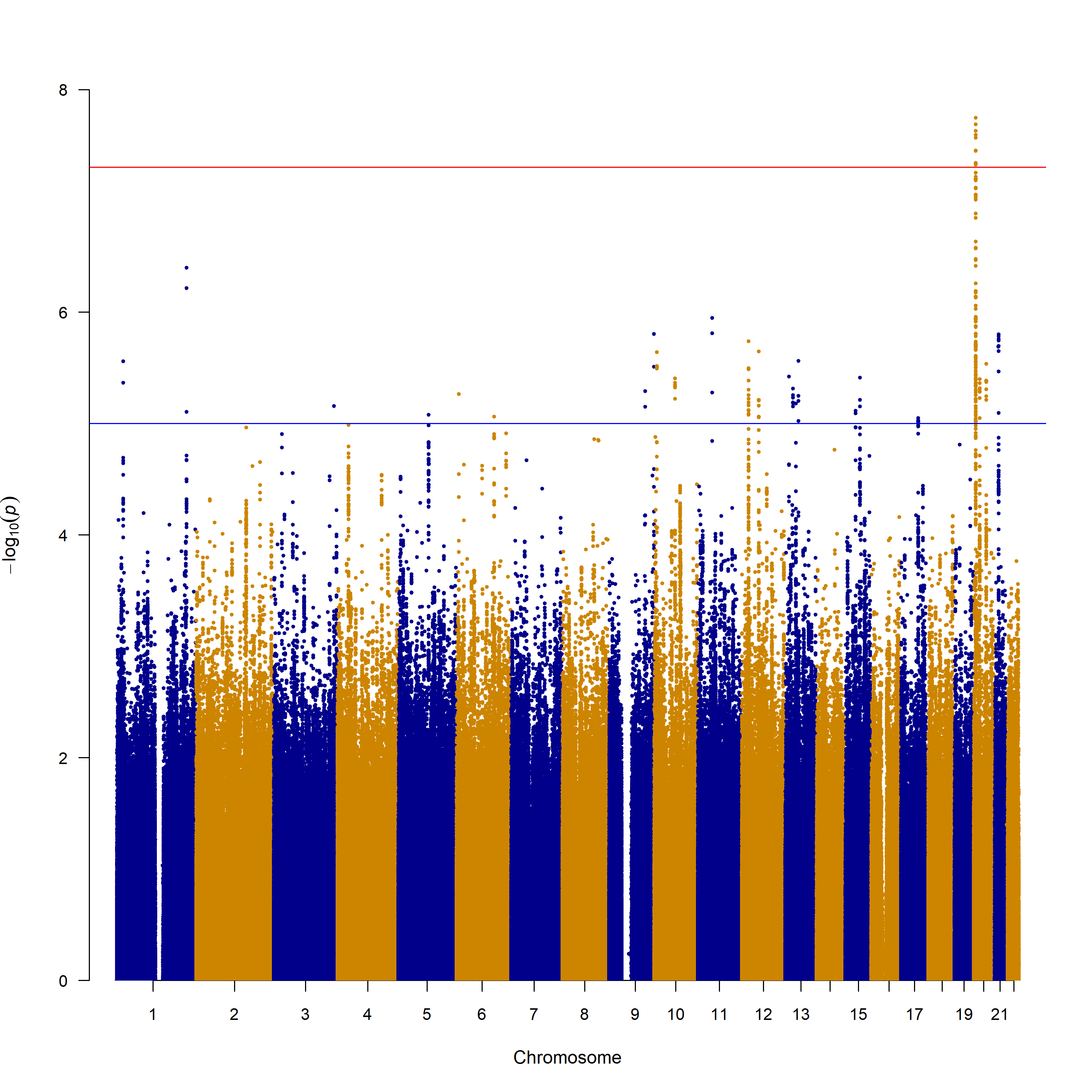

Supplementary Figure 13– Quantile-Quantile plot – Cleft palate only

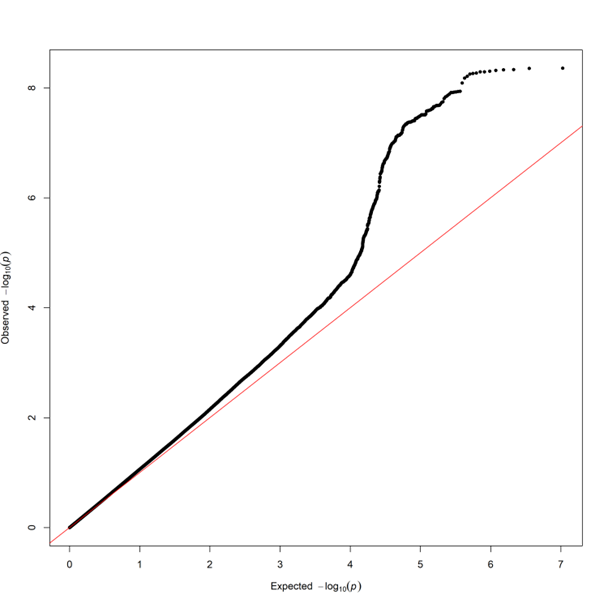

Supplementary Figure 14 – Manhattan plot – Pierre Robin Sequence

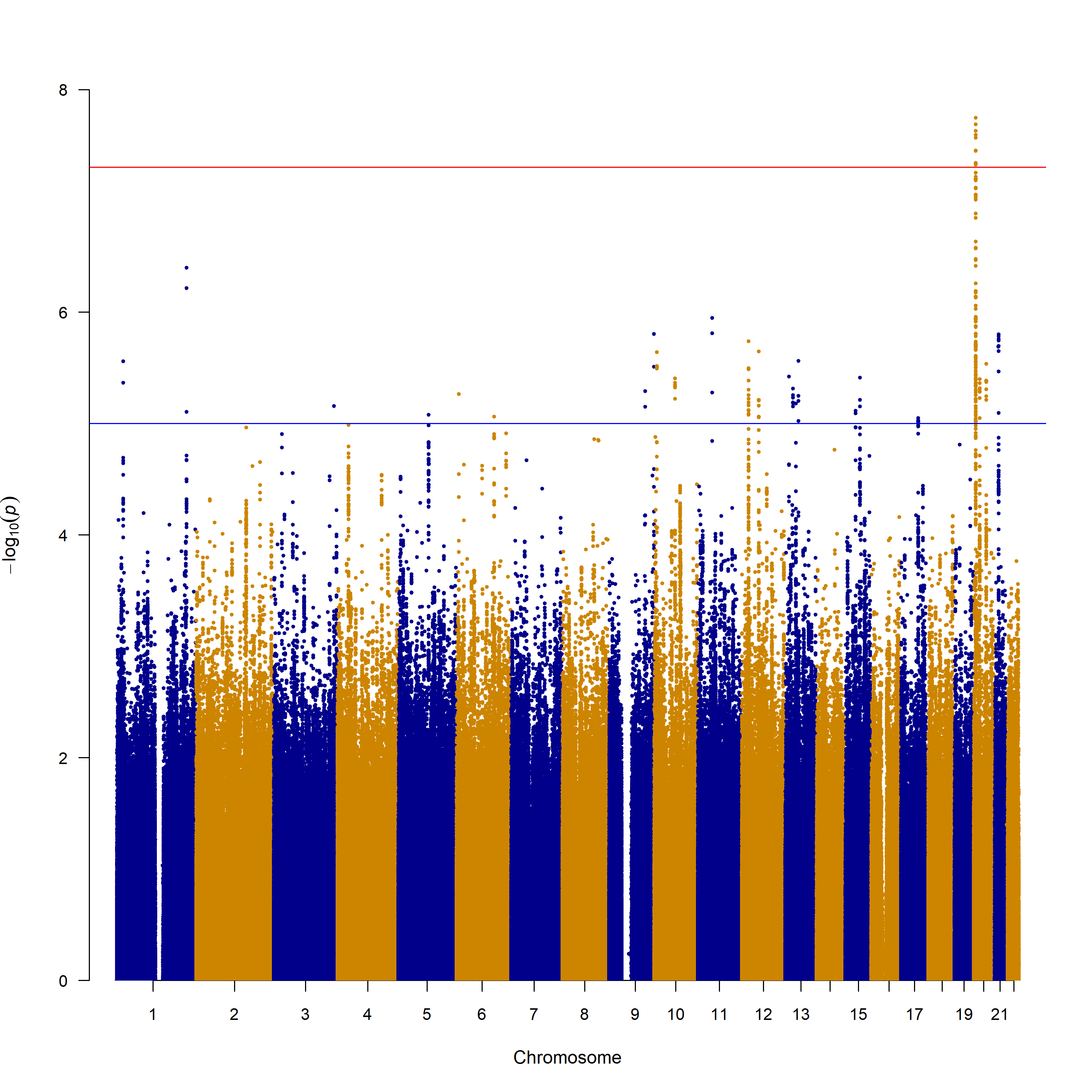

Supplementary Figure 15 –Quantile- Quantile plot – Pierre Robin Sequence

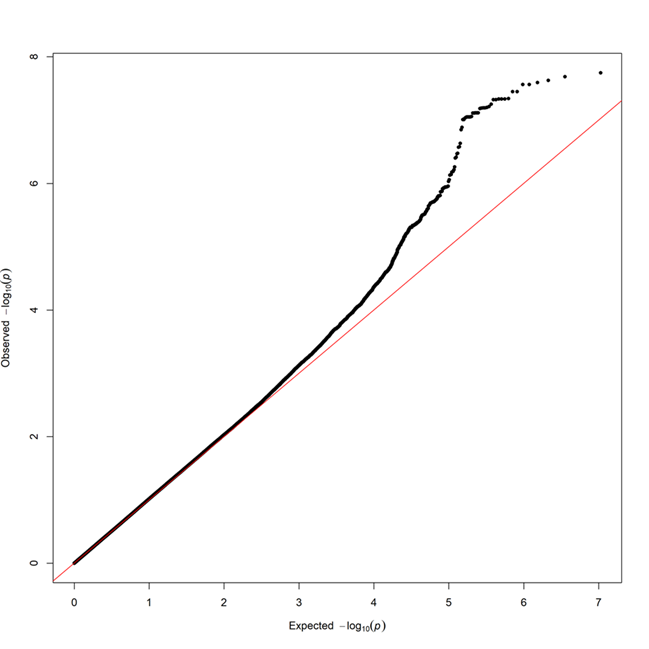

Supplementary Figure 16 – Manhattan plot – Non syndromic cleft palate only without PRS
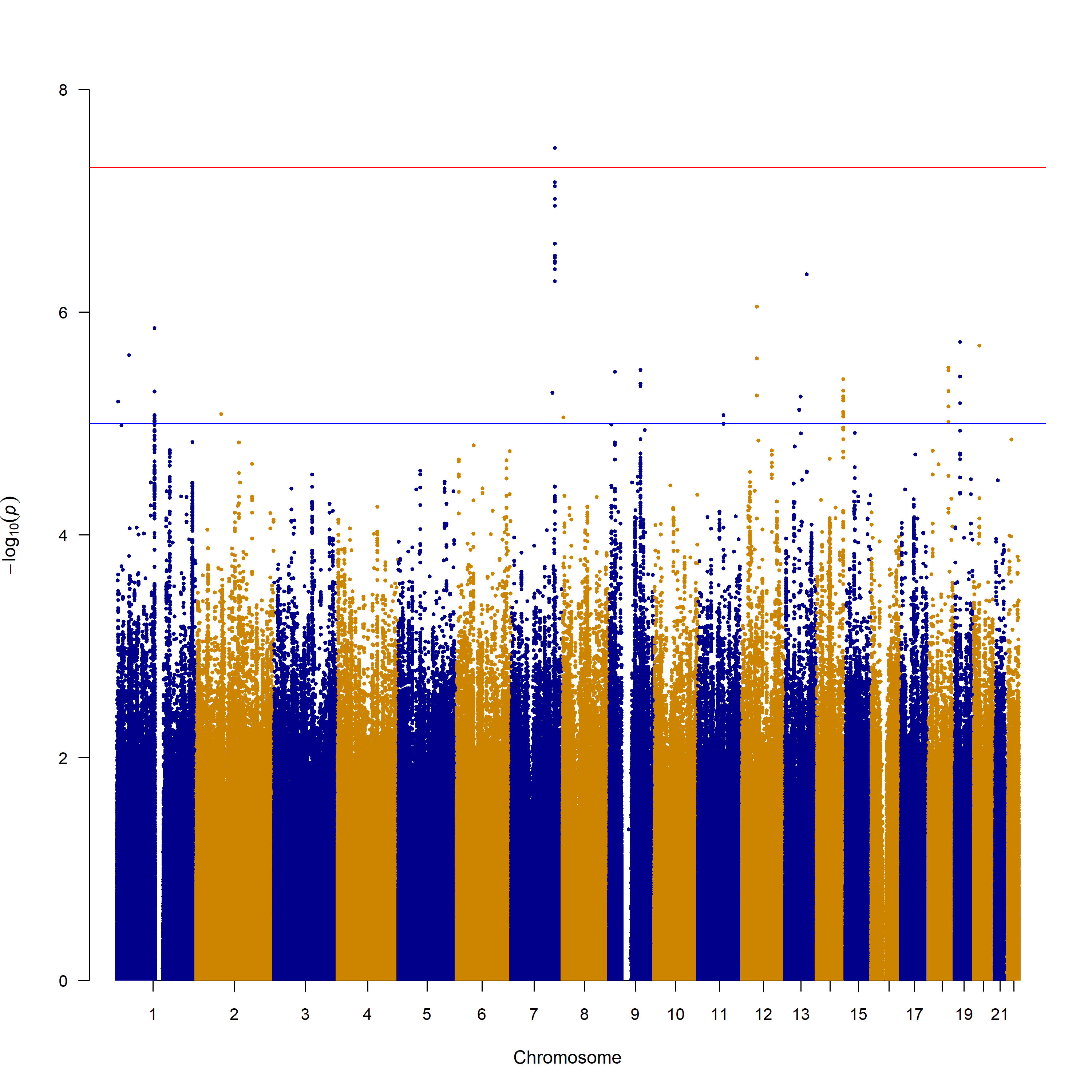

Supplementary Figure 17 – Quantile-Quantile plot – Non syndromic cleft palate only without PRS

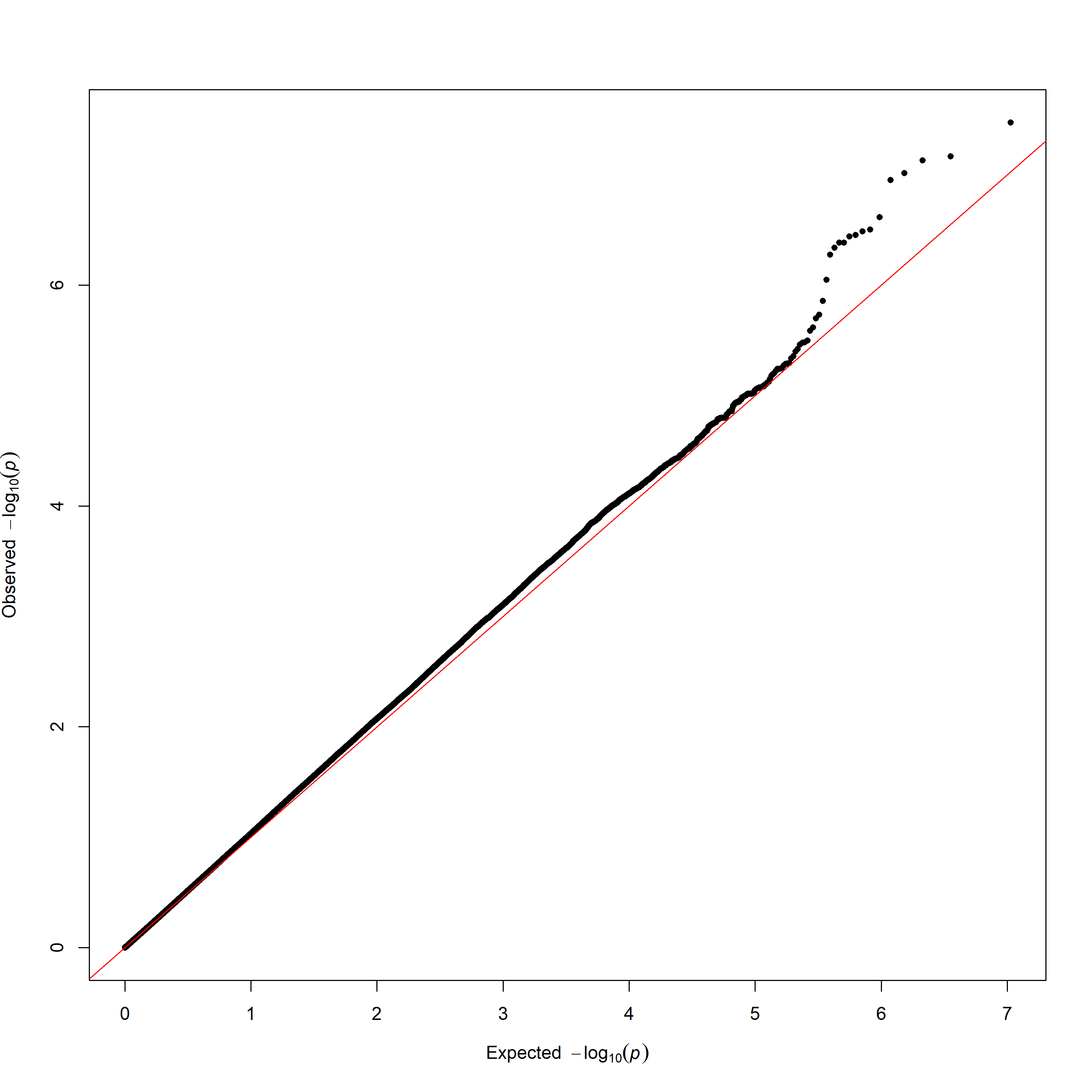
