## Supplementary methods for "Genetic Heterogeneity and Homogeneity Among Orofacial Cleft Subtypes: Genome-Wide Association Studies in the Cleft Collective"

**Sample collection and DNA extraction**

*DNA extraction from blood*

Blood samples were collected from cleft cases by the medical team prior to cleft surgery, and stored in ethylenediaminetetraacetic acid disodium salt (EDTA) tubes to prevent coagulation. DNA was extracted from blood using an in-house guanidine hydrochloride extraction protocol.

*Collection of and extraction of DNA from saliva samples*

For those children recruited after birth, all Cleft Collective parents and controls, saliva samples were self-collected using Oragene 500 kits (<http://www.dnagenotek.com/ROW/products/OG500.html>) which are specifically designed for the collection, preservation, transportation, and purification of DNA from saliva.

**Quantification of DNA**

For all DNA samples, the total quantity of DNA extracted was measured by fluorometric assays using Picogreen reagent. Assays were performed using a Tecan Freedom Evo liquid handling robot and read on an Infinite F2000 Proreader. Samples with a DNA concentration below 40ng/ul were concentrated before use by lyophilisation followed by resuspension in appropriate volume of sterile water.

Fitzsimons, E., Moulton, V., Hughes, D. A., Neaves, S., Ho, K., Hemani, G., Timpson, N., Calderwood, L., Gilbert, E., & Ring, S. (2022). Collection of genetic data at scale for a nationally representative population: The UK Millennium Cohort Study. *Longitudinal and Life Course Studies*, *13*(1), 169–187. <https://doi.org/10.1332/175795921X16334229321103>

Taliun, D., Harris, D. N., Kessler, M. D., et al. (2021). Sequencing of 53,831 diverse genomes from the NHLBI TOPMed Program. Nature, 590, 290–299. https://doi.org/10.1038/s41586-021-03205-y
